# Errors, Hallucinations, and Clinical Impact of General-Purpose Multimodal Large Language Models in Histopathology

**DOI:** 10.64898/2026.09.18.26363369

**Authors:** Kris Lami, Shipra Agarwal, Aleksandra Asaturova, Serdar Balci, Agnes Harahap, Haeyoun Kang, Jennifer Kim, Mina Komuta, Thiyaphat Laohawetwanit, Santosh Menon, Jijgee Munkhdelger, Hoa Hoang Ngoc Pham, Daniel Gomes Pinto, Ayushi Sahay, Swati Satturwar, Kurumi Seki, Maher Sughayer, Yuri Tachibana, Ilknur Turkmen, Qingqing Wu, Ehsan Ullah, Andrey Bychkov, Junya Fukuoka

**Affiliations:** Department of Pathology Informatics, Nagasaki University Graduate School of Biomedical Sciences, Nagasaki, Japan; Department of Pathology, All India Institute of Medical Sciences, India; FSBI «National Medical Research Center for Obstetrics, Gynecology and Perinatology named after Academician V.I.Kulakov» Ministry of Health of the Russian Federation Pirogov Russian National Research Medical University, Moscow, Russia; Pathology Laboratory, Memorial Healthcare Group, Istanbul, Turkey; Department of Anatomical Pathology, Faculty of Medicine, Universitas Indonesia/Dr. Cipto Mangunkusumo Hospital, Jakarta, Indonesia; Department of Pathology, CHA Bundang Medical Center, CHA University School of Medicine, Seongnam, Republic of Korea; Department of Tissue Pathology & Diagnostic Oncology, ICPMR, Westmead Hospital, New South Wales Health Pathology, Sydney, Australia; Department of Pathology, International University of Health and Welfare, School of Medicine, Narita Hospital, Chiba, Japan; Division of Pathology, Chulabhorn International College of Medicine, Thammasat University, Pathum Thani, Thailand; Department of Pathology, Urology and Gynecology Disease Management Group, Homi Bhabha National Institute, Tata Memorial Centre, Mumbai, India; Department of Pathology, Kameda Medical Center, Kamogawa, Japan; Department of Pathology, VNU University of Medicine & Pharmacy, Hanoi, Vietnam; Pathology Department, Hospital Garcia de Orta, Almada, Portugal; Department of Pathology, Tata Memorial Hospital, Tata Memorial Centre, Mumbai, India; Department of Pathology, Wexner Medical Center, The Ohio State University, USA; Department of Pathology and Laboratory Medicine, King Hussein Cancer Center, Amman, Jordan; Department of Surgery, Health New Zealand, Auckland, New Zealand

## Abstract

**Background:** General-purpose large language models (LLMs) are increasingly evaluated in diagnostic pathology, but prior studies have largely emphasized diagnostic accuracy rather than how models fail. We evaluated four LLMs for diagnostic performance, pathology-relevant errors and hallucinations, their burden, and potential clinical impact across multi-organ pathology cases.

**Design:** In this retrospective multicenter study, 153 pathology cases from two institutions spanning 20 organs were evaluated using ChatGPT-5.3 (LLM1), Gemini 3 (LLM2), Grok 4.20 (LLM3), and Claude Opus 4.6 (LLM4). Each LLM received multi-magnification histologic images with clinical context and generated a microscopic description and diagnosis. No data splitting, training, or fine-tuning was performed. Twenty-one pathologists assessed 612 outputs for diagnostic correctness, error and hallucination type and burden, clinical impact (0–4), and overall performance (1–5).

**Results:** Strict diagnostic accuracy was 48.9% overall (60.1% including partially correct diagnoses) and ranged from 36.6% to 58.2% across LLMs. Errors occurred in 82.0% of outputs and hallucinations in 76.6%; 90.2% contained at least one error or hallucination. Misinterpretation was the most frequent error (72.1%), while fabricated histologic features were the dominant hallucination type (74.5%). LLM2 had significantly lower misinterpretation rates than the other three LLMs, while LLM3 had significantly higher fabricated-feature hallucination rates than LLM1 and LLM2. Errors were more frequent in incorrect than correct diagnoses (97.5% vs 66.2%), as were hallucinations (95.1% vs 59.9%; both p < 0.001). Every strictly incorrect diagnosis contained an error and/or hallucination, while 79.9% of strictly correct diagnoses also contained at least one. In multivariable analysis, LLM3 was independently associated with higher odds of an incorrect diagnosis. Diagnostic correctness was the strongest determinant of high clinical impact; each one-point increase in hallucination burden increased the odds by 63% (adjusted OR, 1.63).

**Conclusion:** Diagnostic accuracy alone substantially underestimates the safety limitations of general-purpose LLMs in histopathology. Errors and hallucinations were common even when the final diagnosis was correct, and their burden was independently associated with clinical impact. Evaluation frameworks should therefore assess both diagnostic correctness and the reliability and potential consequences of accompanying generated content.

## Introduction

Large language models (LLMs), initially developed primarily for language understanding and generation, have rapidly evolved into multimodal systems capable of jointly processing textual and visual information. Recent multimodal LLMs integrate pretrained language models with visual encoders, enabling tasks such as image interpretation, visual question answering, and image-grounded conversational reasoning^1–3^. In medicine, multimodal LLMs are increasingly explored for diagnostic support, report generation, education, and clinical decision support^4–6^. By integrating medical images with clinical context and natural-language instructions, these models can perform tasks that more closely resemble multimodal clinical reasoning than text-only LLMs. Early studies have demonstrated their potential across several imaging-intensive specialties, including radiology, ophthalmology, dermatology, and pathology^7–10^. Histopathology represents a particularly relevant application because diagnosis requires integration of microscopic morphology with clinical context, a process these models can now attempt directly from histologic images. Moreover, pathologists are widely using LLMs to support their professional activities^11^. In this setting, multimodal LLMs may be best positioned as assistive tools for pathologists during the microscopic interpretation of tissue specimens from patients undergoing histopathologic evaluation, rather than as autonomous diagnostic systems, with final diagnostic responsibility remaining with the pathologist.^12,13^

Several studies have evaluated the performance of LLMs in the field of pathology. A pathology-specific model, PathChat, achieved 78.1% accuracy using histology images alone and 89.5% when clinical context was added in a multi-organ diagnostic benchmark, illustrating the potential value of multimodal pathology models^10^. Evaluations of general-purpose multimodal LLMs across histopathology and cytology have shown highly variable diagnostic performance, influenced by case complexity, organ system, task formulation, and prompting strategy. Collectively, these studies suggest that although such models can extract diagnostically relevant morphologic information, their performance remains inconsistent for fine-grained classification and specific diagnoses^14–16^.

A fundamental limitation of generative LLMs is their ability to produce fluent and plausible responses that nevertheless contain incorrect, unsupported, or internally inconsistent information. Such failures may take several forms, including omission of relevant findings, misinterpretation of genuine features, fabrication of findings that are not present, unsupported diagnostic or clinical inferences, contradictions, and failure to follow instructions^17–21^. In healthcare, these errors are particularly concerning because outputs may appear authoritative even when portions of the generated evidence or reasoning are unreliable. Studies in other medical applications have shown that hallucinations may occur despite otherwise acceptable task performance and can have direct safety implications. In clinical summarization, 44% of identified hallucinations were classified as major errors with potential consequences for diagnosis or management, while a diagnostic study demonstrated substantial variation in hallucination rates across models despite near-perfect diagnostic accuracy^22,23^. These findings suggest that correctness of the final diagnosis does not necessarily guarantee the reliability of the information or reasoning generated to support it.

Despite growing interest in image-capable LLMs for diagnostic pathology, most evaluations have focused primarily on diagnostic or classification accuracy, with limited assessment of the quality and reliability of generated microscopic interpretations. Recent work has begun to move beyond accuracy alone; for example, expert pathologists have evaluated diagnostic reasoning across multiple LLMs and identified differences in coherence, analytical depth, and pathology-specific reasoning strategies. However, this evaluation was text-based and did not assess image-grounded failures^24^. Consequently, it remains unclear how often clinically relevant errors and hallucinations occur in multimodal pathology outputs, whether they persist despite a correct diagnosis, and whether their burden and potential impact vary across models and case characteristics. This distinction is clinically important because outputs with similar diagnostic accuracy may have substantially different safety profiles, meaning that accuracy alone may underestimate clinically relevant limitations.

To address this gap, we systematically evaluated four contemporary general-purpose multimodal LLMs across multi-organ histopathology cases. Beyond diagnostic performance, we characterized pathology-relevant errors and hallucinations, quantified their burden, and estimated their potential clinical impact. We further examined differences across models, organs/systems, disease categories, rarity, and diagnostic efficiency, as well as the relationships between error and hallucination burden, diagnostic correctness, and overall performance. As an evaluation study, no model development, training, or fine-tuning was performed.

## Results

### Dataset characteristics

An overview of the study design is provided in Figure 1. A total of 153 cases spanning 20 organs/systems were included, with no duplicate consensus diagnoses within the same organ/system (Supplementary Table 1). Four general-purpose LLMs were evaluated: ChatGPT-5.3 (OpenAI, San Francisco, CA, USA; LLM1), Gemini 3 (Google DeepMind, London, UK; LLM2), Grok 4.20 (SpaceXAI, Palo Alto, CA, USA; LLM3), and Claude Opus 4.6 (Anthropic, San Francisco, CA, USA; LLM4). Each model evaluated all 153 cases, generating 612 outputs in total. The dataset comprised 122 neoplastic and 31 non-neoplastic cases; 67 common, 63 sporadic, and 23 rare diseases; and 55 cases classified as fully sufficient for diagnosis, 40 in which additional support was desirable, and 58 in which additional support was required.

**Figure 1.**
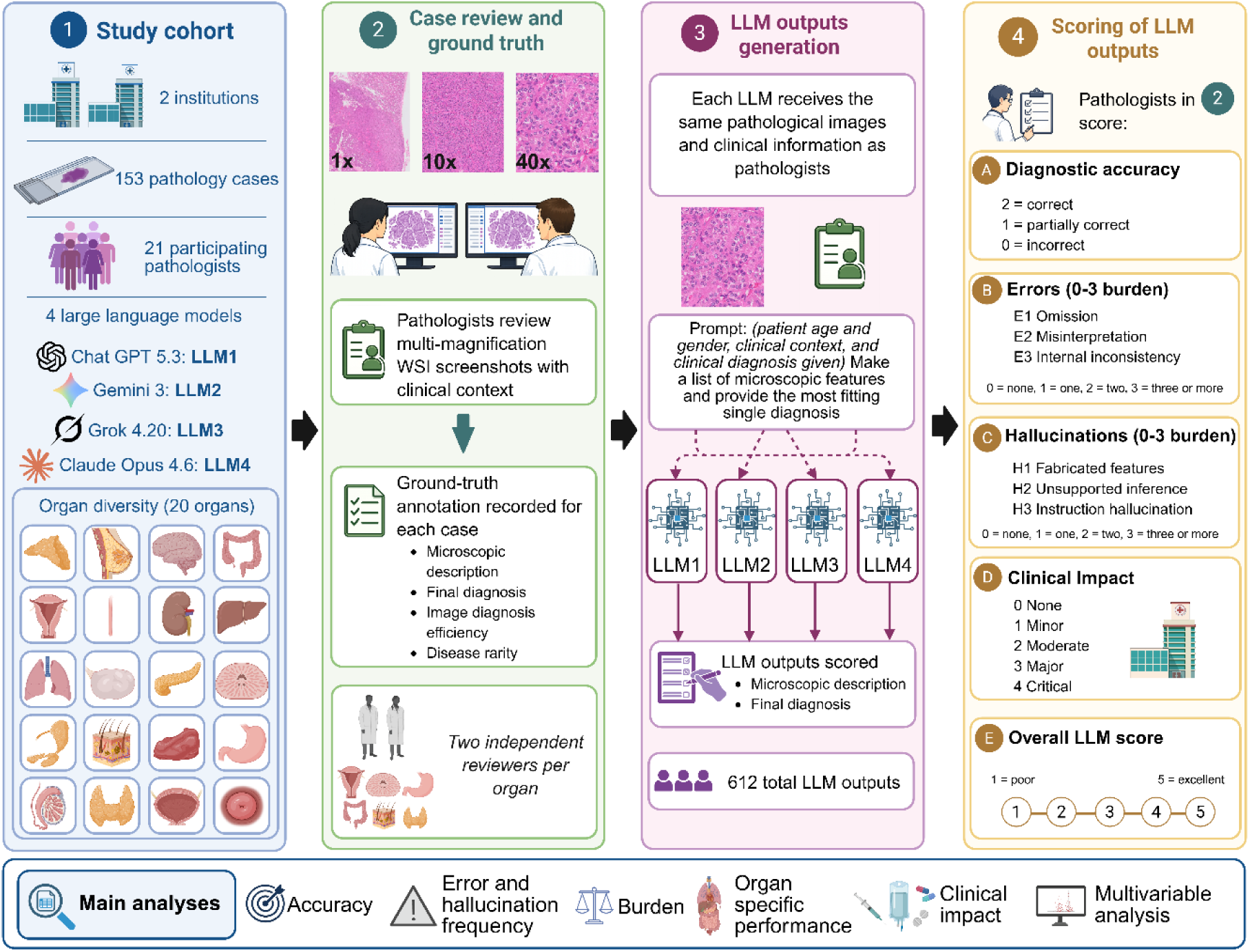
Overview of the study design and evaluation framework. The study cohort comprised 153 histopathology cases from two institutions, spanning 20 organs/systems, with evaluation involving 21 pathologists and four general-purpose multimodal LLMs: ChatGPT-5.3 (LLM1), Gemini 3 (LLM2), Grok 4.20 (LLM3), and Claude Opus 4.6 (LLM4). In step 1, the study cohort and participating models are summarized. In step 2, pathologists reviewed multi-magnification histologic images with clinical context and established the reference diagnosis and case-level annotations, including microscopic description, diagnostic efficiency, and disease rarity. Two pathologists independently reviewed each case. In step 3, each LLM received the same histologic images and clinical information and was prompted to generate a microscopic description and a single most likely diagnosis, yielding 612 LLM outputs. In step 4, blinded pathologists evaluated the outputs for diagnostic correctness, pathology-relevant errors, hallucinations, clinical impact, and overall performance. Diagnostic correctness was scored as correct, partially correct, or incorrect. Error and hallucination subtypes were scored from 0 to 3 according to the number of occurrences, clinical impact from 0 to 4 according to potential clinical consequences, and overall LLM performance from 1 to 5. The principal analyses included diagnostic accuracy, error and hallucination frequency, cumulative burden, organ/system-specific performance, clinical impact, and multivariable analyses of adverse outcomes. LLM, large language model; WSI, whole-slide image. Created with BioRender.com.

### Diagnostic performance across models and case characteristics

Overall strict diagnostic accuracy was 48.9%, increasing to 60.1% when partially correct diagnoses were included. Performance varied across models: strict accuracy was 47.1%, 53.6%, 36.6%, and 58.2% for LLM1–4, respectively, while partial accuracy increased to 63.4%, 64.7%, 45.8%, and 66.7%. LLM3 showed the lowest performance and was significantly less accurate than LLM2 and LLM4 (Figure 2A). At the case level, at least one LLM provided a strictly correct diagnosis in 118/153 cases (77.1%), whereas all four were correct in only 29 cases (19.0%) and none were correct in 22 (14.4%). In 23 cases (15.0%), only one model reached the correct diagnosis, most often LLM4 (8 cases), followed by LLM2 (7), LLM1 (5), and LLM3 (3), suggesting some complementary performance across models (Supplementary Table 2). Accuracy was higher for neoplastic than for non-neoplastic cases (52.5% vs 34.7%, *p* = 0.00042), although this difference was significant individually only for LLM4 (*p* = 0.029; Figure 2B). In contrast, disease rarity had little influence on performance, with strict accuracy of 52.2%, 46.0%, and 46.7% for common, sporadic, and rare diseases, respectively, without significant differences overall or by LLM (Figure 2C). Diagnostic efficiency had a clearer effect: accuracy was 53.6% for fully sufficient cases, 57.5% when additional support was desirable, and 38.4% when support was required, with the latter significantly lower than the other groups (*p* = 0.0002); no per-LLM comparison remained significant after correction (Figure 2D). Performance also varied substantially by organ/system, with the highest accuracy in uterine cervix, pancreas, CNS, breast, skin, and urinary bladder, and the lowest in lung, colon, prostate, stomach, testis, and thyroid (Supplementary Tables 3 and 4).

**Figure 2.**
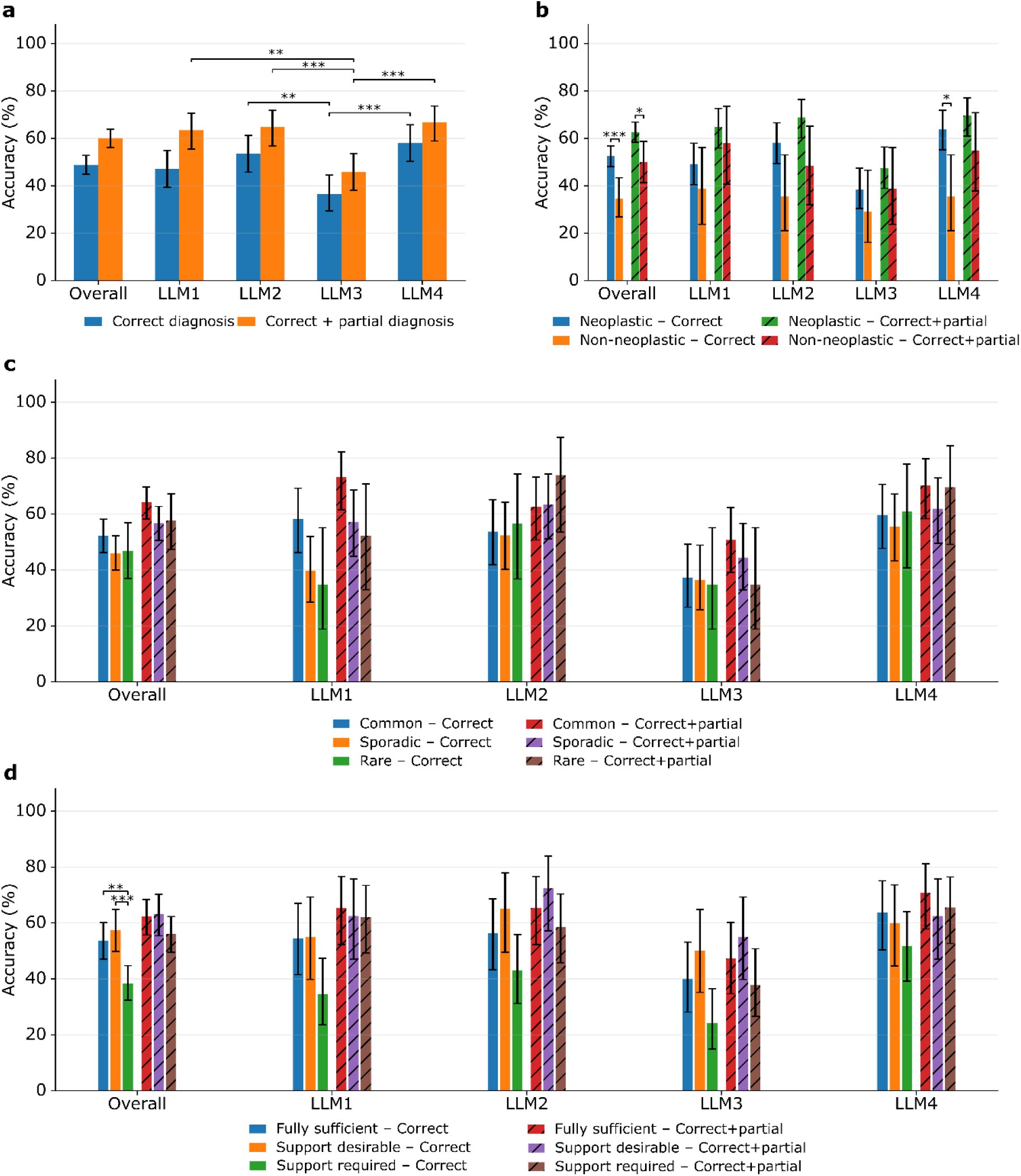
Diagnostic accuracy of multimodal large language models across case characteristics. **a,** Overall strict diagnostic accuracy (correct diagnosis) and partial diagnostic accuracy (correct or partially correct diagnosis), shown for all outputs combined and for each LLM. **b,** Diagnostic accuracy according to disease category (neoplastic versus non-neoplastic), overall and by LLM. **c,** Diagnostic accuracy according to disease rarity (common, sporadic, and rare), overall and by LLM. **d,** Diagnostic accuracy according to diagnostic efficiency of the provided material (fully sufficient, additional support desirable, or additional support required), overall and by LLM. Solid bars represent strict diagnostic accuracy, whereas hatched bars represent accuracy including partially correct diagnoses; colors distinguish the case categories within each panel as indicated in the legends. Error bars represent 95% Wilson binomial confidence intervals. Brackets indicate statistically significant pairwise comparisons based on paired tests accounting for the matched case-level design; one asterisk indicates *p* < 0.05, two asterisks *p* < 0.01, and three asterisks *p* < 0.001. Only statistically significant comparisons are shown. LLM, large language model.

### Patterns of errors and hallucinations across models and case characteristics

Errors were classified as omission, misinterpretation, or internal inconsistency, whereas hallucinations were classified as fabricated feature, unsupported inference, or instruction hallucination (Supplementary Figure 1). Errors and hallucinations were highly prevalent across the 612 LLM outputs: 502 outputs (82.0%) contained at least one error, 469 (76.6%) contained at least one hallucination, and 552 (90.2%) contained either. Misinterpretation was the dominant error type, occurring in 441 outputs (72.1%), followed by omission in 326 (53.3%) and internal inconsistency in 105 (17.2%). Fabricated histologic features were the most frequent hallucination type, occurring in 456 outputs (74.5%), whereas unsupported inference and instruction hallucination occurred in 140 (22.9%) and 77 (12.6%), respectively (Figure 3A). Error patterns differed across models: LLM2 had the lowest overall error rate (74.5%), whereas LLM3 had the highest error and hallucination rates (88.2% and 83.7%, respectively); LLM1 and LLM2 shared the lowest hallucination rate (72.5%; Figure 3B). Misinterpretation remained the most frequent error for every model but was significantly less common with LLM2 than with LLM1, LLM3, and LLM4 (all Holm-adjusted *p* < 0.05). Fabricated features similarly predominated across models and were significantly more frequent with LLM3 than with LLM1 and LLM2 (all *p* < 0.05; Figure 3C). Error and hallucination rates were comparable between neoplastic and non-neoplastic diseases (Supplementary Figure 2), whereas both increased numerically with disease rarity: compared with common diseases, rare diseases showed 9.4% higher error rates (87.0% vs 77.6%) and 11.3% higher hallucination rates (82.6% vs 71.3%). This pattern was most apparent for LLM1, although no per-LLM comparison across rarity categories remained significant after multiple-testing correction (Supplementary Figure 3). Diagnostic efficiency showed a different pattern: error rates remained relatively stable across categories (80.9% in fully sufficient cases vs 83.6% when additional support was required), whereas hallucination rates increased more clearly with the need for additional diagnostic support (71.8% vs 81.5%), particularly for LLM1, LLM2, and LLM4 (Supplementary Figure 4). Detailed subtype distributions by disease category, rarity, and diagnostic efficiency are provided in Supplementary Tables 5–7. The strongest relationship emerged with diagnostic correctness. Errors were present in 97.5% of incorrect versus 66.2% of correct diagnoses, and hallucinations in 95.1% versus 59.9%, respectively, with similar trends across all four models (Figure 3D). Notably, every strictly incorrect diagnosis contained at least one error or hallucination, whereas 239 of 299 strictly correct diagnoses (79.9%) also contained at least one, indicating that a correct final diagnosis did not necessarily imply a reliable accompanying interpretation (Supplementary Table 8). Among outputs containing errors or hallucinations, LLM4 had higher adjusted odds of being diagnostically correct than LLM3 (OR 2.27, 95% CI 1.51–3.43, *p* < 0.001). Conversely, LLM3 had higher adjusted odds of being diagnostically incorrect than LLM1 (OR 1.85, 95% CI 1.22–2.82), LLM2 (OR 2.20, 95% CI 1.50–3.22), and LLM4 (OR 2.51, 95% CI 1.66–3.81; all *p* < 0.05). Error and hallucination rates also varied substantially across organs/systems, with LLM3 generally showing higher rates and LLM2 comparatively lower error rates, although these patterns were not uniform across all organ systems (Supplementary Tables 9–13).

**Figure 3.**
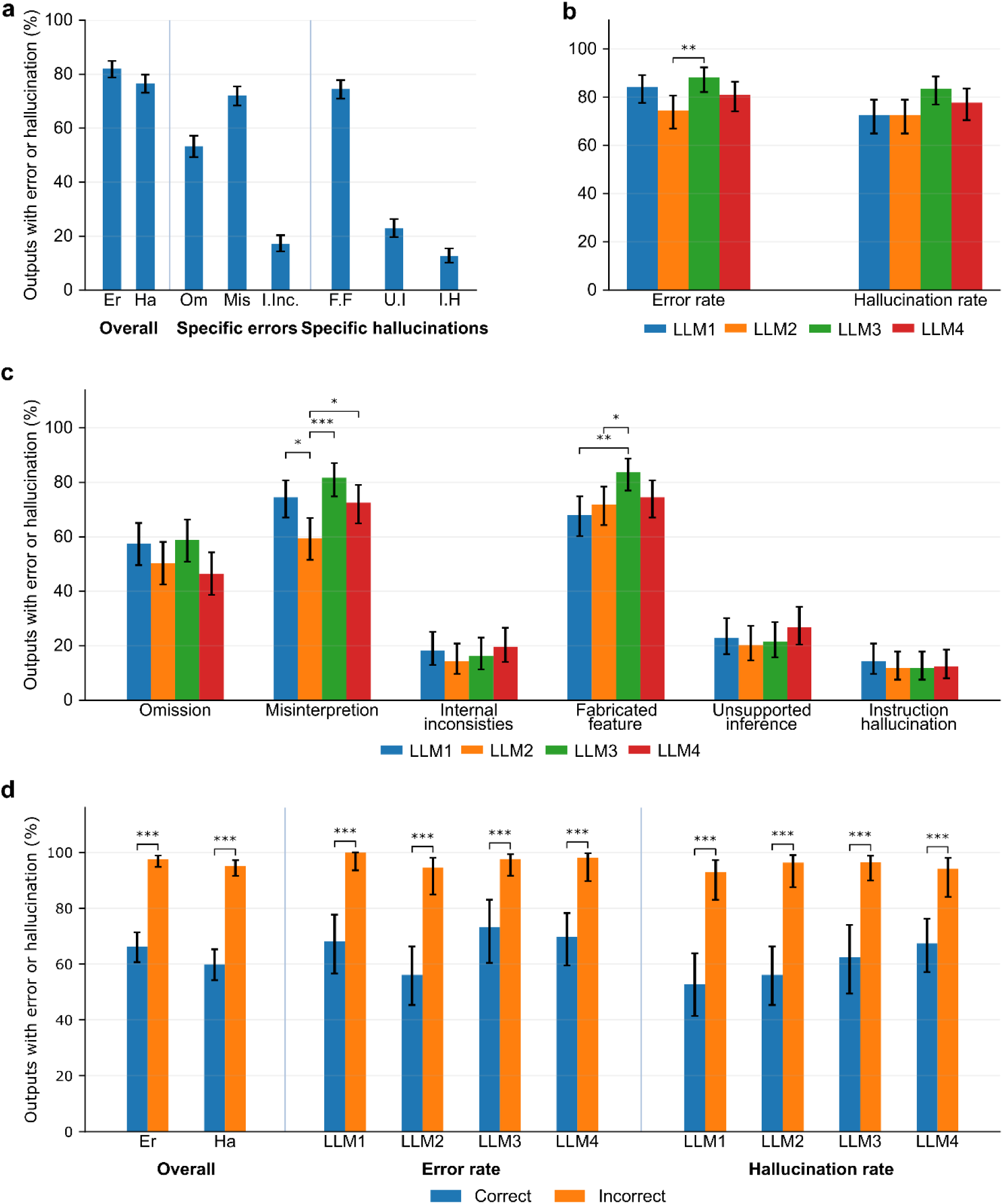
Patterns of errors and hallucinations across multimodal large language models and diagnostic correctness. **a,** Overall prevalence of errors and hallucinations and of their specific subtypes across all LLM outputs. **b,** Overall error and hallucination rates for each LLM. **c,** Prevalence of individual error and hallucination subtypes for each LLM. **d,** Overall and model-specific error and hallucination rates according to strict diagnostic correctness. Partially correct diagnoses are not included in panel d. Error bars represent 95% Wilson binomial confidence intervals. Brackets indicate statistically significant comparisons; one asterisk indicates *p* < 0.05, two asterisks *p* < 0.01, and three asterisks *p* < 0.001. Only statistically significant comparisons are shown. Er, error; Ha, hallucination; Om, omission; Mis, misinterpretation; I.Inc., internal inconsistency; F.F, fabricated feature; U.I, unsupported inference; I.H, instruction hallucination; LLM, large language model.

### Error and hallucination burden across models and case characteristics

Error and hallucination burden scores ranged from 0 to 9, reflecting the summed 0–3 scores across the three respective subtypes. Mean burden differed significantly among the four LLMs, with LLM3 showing the highest error burden (3.17 ± 2.16, 95% CI 2.83–3.51) and hallucination burden (2.63 ± 1.95, 95% CI 2.32–2.94). Its error burden was significantly higher than that of LLM2 (2.16 ± 2.05, 95% CI 1.83–2.48; *p* < 0.001) and LLM4 (2.52 ± 2.14, 95% CI 2.18–2.86; *p* = 0.008), while its hallucination burden was significantly higher than that of LLM1 (1.99 ± 1.84, 95% CI 1.70–2.29; *p* < 0.001) and LLM2 (2.00 ± 1.90, 95% CI 1.70–2.30; *p* < 0.001). LLM2 also had a significantly lower error burden than LLM1 (2.16 ± 2.05 vs 2.75 ± 2.08; *p* = 0.021; Figure 4A). By disease category, error burden was similar between neoplastic and non-neoplastic cases, whereas hallucination burden was higher in non-neoplastic cases, although this difference was not significant (*p* = 0.095; Figure 4B). Neither disease rarity nor diagnostic efficiency was significantly associated with burden, although rare diseases showed the highest numerical hallucination burden and hallucination burden increased from fully sufficient to support-required cases (Figures 4C and 4D). The sharpest differences emerged with diagnostic correctness: error burden was approximately 2.9 points higher in incorrect than correct diagnoses (4.17 ± 1.98, 95% CI 3.84–4.51 vs 1.28 ± 1.25, 95% CI 1.11–1.46; *p* < 0.001), while hallucination burden was approximately 1.9 points higher (3.16 ± 1.82, 95% CI 2.84–3.48 vs 1.29 ± 1.44, 95% CI 1.06–1.52; *p* < 0.001; Figure 4E). Burden also varied across organs/systems, further indicating that the accumulation of model failures depended strongly on case context (Supplementary Table 14).

**Figure 4.**
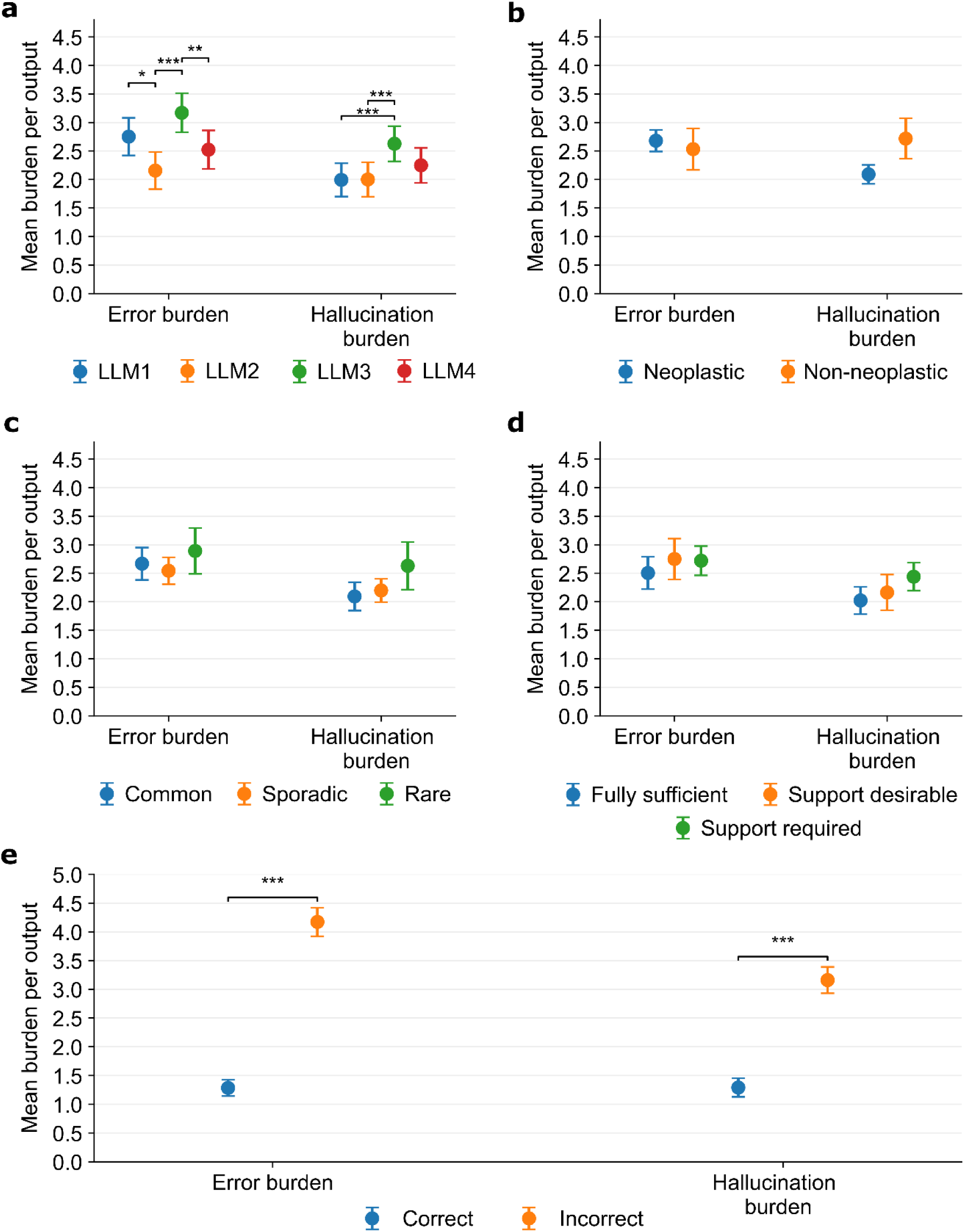
Error and hallucination burden across multimodal large language models and case characteristics. **a,** Mean error and hallucination burden for each LLM. **b,** Mean burden according to disease category (neoplastic versus non-neoplastic). **c,** Mean burden according to disease rarity (common, sporadic, and rare). **d,** Mean burden according to diagnostic efficiency of the provided material (fully sufficient, additional support desirable, or additional support required). **e,** Mean error and hallucination burden according to strict diagnostic correctness. Partially correct diagnoses are not included in panel e. Error burden represents the summed scores for omission, misinterpretation, and internal inconsistency, and hallucination burden represents the summed scores for fabricated features, unsupported inference, and instruction hallucination, each ranging from 0 to 9 per output. Points represent mean burden per output and error bars represent 95% confidence intervals. Brackets indicate statistically significant comparisons; one asterisk indicates *p* < 0.05, two asterisks *p* < 0.01, and three asterisks *p* < 0.001. Only statistically significant comparisons are shown. LLM, large language model.

### Clinical impact and safety profile of LLM outputs

Clinical impact was scored from 0 to 4, with higher scores indicating greater potential harm if the LLM output were adopted in clinical practice. The overall clinical impact score was 1.78 ± 1.53 (95% CI 1.66–1.90) and differed significantly among models (Friedman χ²[3] = 26.26, *p* < 0.001). LLM3 had the highest mean clinical impact score (2.21 ± 1.56, 95% CI 1.96– 2.46), compared with LLM1 (1.76 ± 1.45, 95% CI 1.53–1.99), LLM2 (1.61 ± 1.54, 95% CI 1.36–1.85), and LLM4 (1.55 ± 1.51, 95% CI 1.31–1.79), whereas no significant differences were observed among LLM1, LLM2, and LLM4 (Figure 5A). Thus, outputs from LLM3 carried a greater potential for clinically consequential effects. Clinical impact was also higher in non-neoplastic than neoplastic cases (2.26 ± 1.22, 95% CI 1.81–2.71 vs 1.66 ± 1.09, 95% CI 1.46–1.86; *p* = 0.010; Figure 5B), suggesting that failures in non-neoplastic conditions may be more likely to influence diagnostic interpretation or management. In contrast, clinical impact did not differ significantly by disease rarity or diagnostic efficiency, despite numerical increases across both categories (Figures 5C and 5D), and varied across organs/systems, with the highest scores in esophageal cases and the lowest in soft-tissue cases (Supplementary Figure 5). To further examine clinical safety, outputs were classified as safe correct when the diagnosis was strictly correct, clinical impact was ≤2, and neither error nor hallucination burden was high (<3), and as dangerous wrong when the diagnosis was incorrect, clinical impact was ≥3, and either error or hallucination burden was high (≥3). Overall, 206/612 outputs (33.7%) were safe correct, representing 68.9% of all strictly correct diagnoses, whereas 175 (28.6%) were dangerous wrong, accounting for 71.7% of all incorrect diagnoses. Safe-correct outputs were more frequent with LLM2 and LLM4 than with LLM3, while dangerous-wrong outputs were significantly more frequent with LLM3 than with LLM1, LLM2, or LLM4 (*p* ≤ 0.001; Figure 5E).

**Figure 5.**
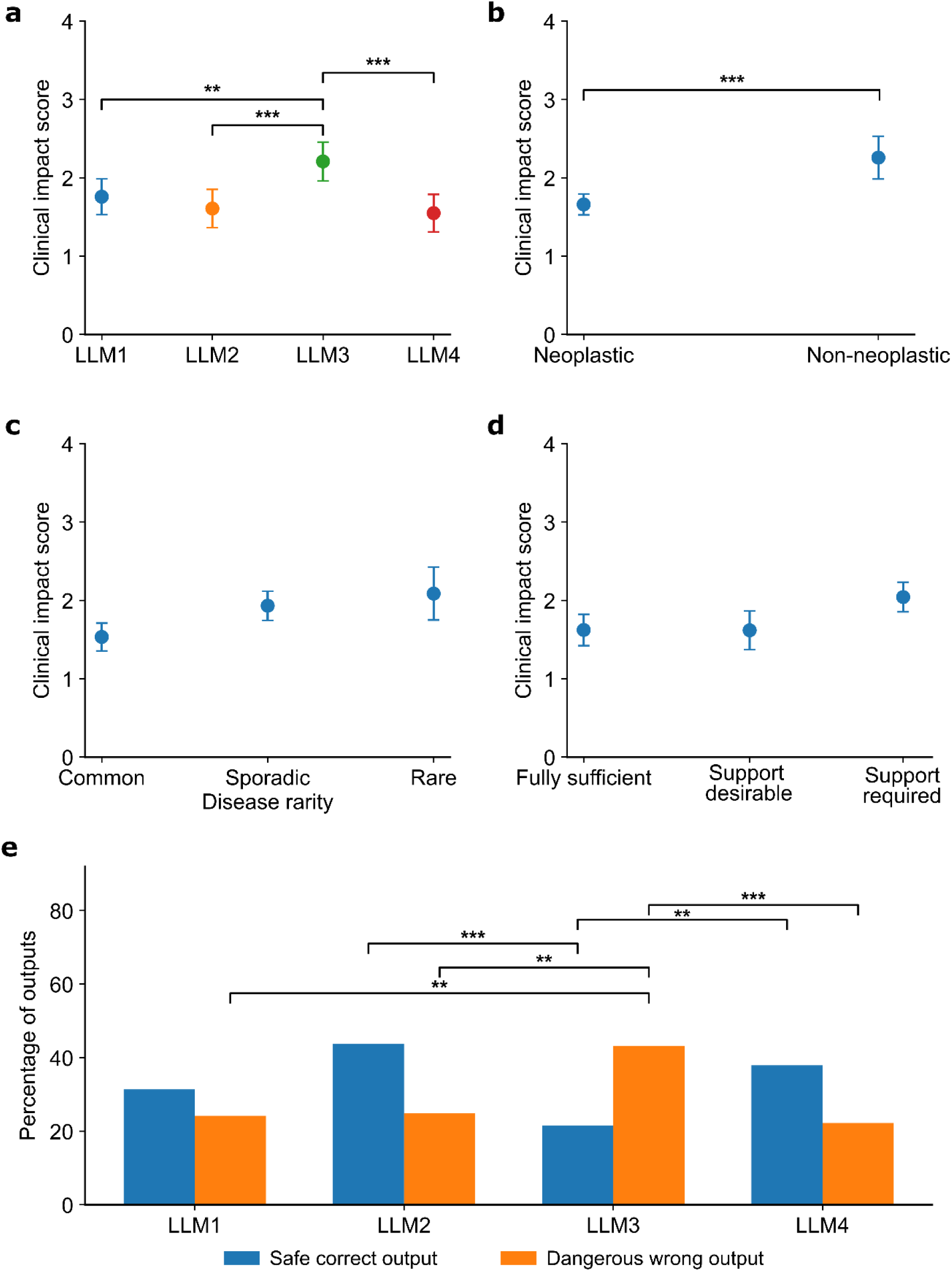
Clinical impact and safety profiles of multimodal large language model outputs. **a,** Mean clinical impact score for each LLM. **b,** Mean clinical impact score according to disease category (neoplastic versus non-neoplastic). **c,** Mean clinical impact score according to disease rarity (common, sporadic, and rare). **d,** Mean clinical impact score according to diagnostic efficiency of the provided material (fully sufficient, additional support desirable, or additional support required). Clinical impact was scored from 0 to 4, with higher scores indicating greater potential clinical consequences. Points represent mean scores and error bars represent 95% confidence intervals around the mean, calculated as the mean plus or minus 1.96 times the standard error. **e,** Proportion of outputs classified as safe correct or dangerous wrong for each LLM. Safe-correct outputs had a strictly correct diagnosis, clinical impact score of 2 or lower, and no high-burden error or hallucination (<3); dangerous-wrong outputs had an incorrect diagnosis, clinical impact score of 3 or higher, and a high error or hallucination burden (≥3). Brackets indicate statistically significant comparisons; two asterisks indicate *p* < 0.01 and three asterisks *p* < 0.001. Only statistically significant comparisons are shown. LLM, large language model.

### Overall LLM performance scores

Overall LLM performance was scored on a 1–5 scale based on the quality of the microscopic description and final diagnosis, incorporating diagnostic accuracy and the associated error and hallucination burden. Across all outputs, the mean overall score was 3.19 ± 1.42 (95% CI 3.08–3.31). LLM4 achieved the highest mean score (3.40 ± 1.40, 95% CI 3.18–3.63), followed closely by LLM2 (3.33 ± 1.48, 95% CI 3.09–3.56) and LLM1 (3.30 ± 1.29, 95% CI 3.10–3.51), whereas LLM3 had the lowest score (2.75 ± 1.44, 95% CI 2.52–2.97). LLM3 scored significantly lower than LLM1, LLM2, and LLM4, while no significant differences were observed among the other three models. Further details, such as overall performance according to disease category, disease rarity, diagnostic efficiency, and organ/system are shown in Supplementary Figure 6.

### Factors independently associated with adverse LLM outcomes

Multivariable analyses identified several factors independently associated with diagnostic inaccuracy, errors, hallucinations, high clinical impact, and low overall LLM performance. LLM identity remained an important determinant of diagnostic accuracy: LLM3 had higher odds of an incorrect diagnosis than LLM1 (adjusted odds ratio [aOR], 2.99; 95% credible interval [CrI], 2.03-4.42; Holm-adjusted *p* < 0.001) and LLM2 (aOR 3.29; 95% CrI, 1.87-5.78; *p* < 0.001), whereas LLM4 had lower odds than LLM3 (aOR, 0.27; 95% CrI, 0.15-0.47; *p* < 0.001); no significant differences were observed among LLM1, LLM2, and LLM4. Independent of model identity, incorrect diagnosis was also associated with non-neoplastic disease (aOR, 2.18; 95% CrI, 1.39-3.41), sporadic disease rarity (aOR, 1.50; 95% CrI, 1.09–2.06), and cases requiring additional diagnostic support (aOR, 1.47; 95% CrI, 1.07–2.03; Figure 6A). For errors, LLM2 had lower adjusted odds than LLM1 (aOR, 0.47; 95% CrI, 0.31–0.70; *p* = 0.002), whereas LLM3 had higher odds than LLM2 (aOR, 3.34; 95% CrI, 1.68–6.61; *p* = 0.003). Sporadic and rare diseases were additionally associated with increased odds of error compared with common diseases (aOR, 1.80 and 2.33, respectively; Figure 6B). Hallucinations showed a similar model-dependent pattern, with LLM3 having higher odds than LLM1 (aOR, 2.30; 95% CrI, 1.44–3.69; *p* = 0.003) and LLM2 (aOR, 2.29; 95% CrI, 1.23–4.25; *p* = 0.043). Beyond LLM identity, hallucinations were associated with sporadic disease rarity (aOR, 1.48; 95% CrI, 1.06–2.08) and with cases in which additional diagnostic support was desirable (aOR, 1.53; 95% CrI, 1.01–2.30) or required (aOR, 1.91; 95% CrI, 1.33–2.75; Figure 6C).

**Figure 6.**
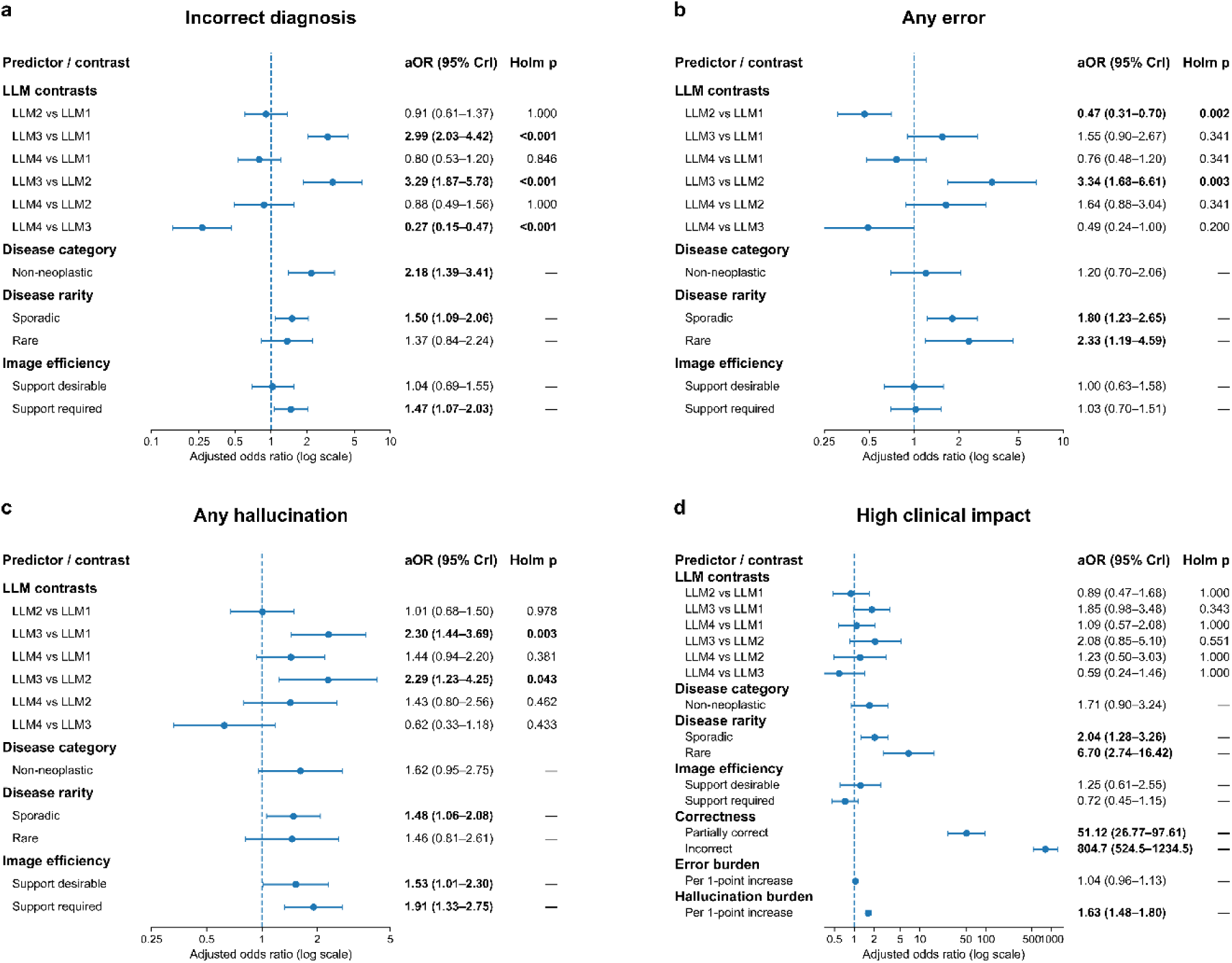
Multivariable analysis of factors independently associated with adverse multimodal large language model outcomes. **a,** Adjusted associations with incorrect diagnosis. **b,** Adjusted associations with the presence of any error. **c,** Adjusted associations with the presence of any hallucination. **d,** Adjusted associations with high clinical impact, defined as a clinical impact score of 3–4. Blue points represent adjusted odds ratios (aORs) and horizontal blue bars represent 95% credible intervals (CrIs); the vertical blue dashed line indicates an aOR of 1. For disease category, neoplastic disease is the reference; for disease rarity, common disease is the reference; and for diagnostic efficiency, fully sufficient material is the reference. In panel d, correct diagnosis is the reference for diagnostic correctness, and error and hallucination burdens are modeled per one-point increase. Model effects are shown as all pairwise LLM contrasts. Values to the right of 1 indicate higher adjusted odds of the corresponding adverse outcome, whereas values to the left indicate lower adjusted odds. Bold estimates denote statistically supported associations; for LLM contrasts, Holm-adjusted *p* values are shown, whereas for other predictors support was based on 95% CrIs excluding 1. LLM, large language model; aOR, adjusted odds ratio; CrI, credible interval.

The strongest associations appeared for clinical impact and overall performance. Compared with correct diagnoses, partially correct diagnoses had markedly higher odds of high clinical impact (aOR, 51.12; 95% CrI, 26.77–97.61), while incorrect diagnoses had an even greater association (aOR, 804.68; 95% CrI, 524.50–1234.52). Sporadic and rare diseases were also independently associated with high clinical impact (aOR 2.04 and 6.70, respectively), and each one-point increase in hallucination burden increased the odds by 63% (aOR, 1.63; 95% CrI, 1.48–1.80; Figure 6D). Similarly, LLM3 had higher odds of a low overall score than LLM1 (aOR, 3.40; 95% CrI, 1.71–6.78; *p* = 0.003). Partially correct and incorrect diagnoses were strongly associated with low scores (aOR 13.37 and 1251.52, respectively), while each one-point increase in error and hallucination burden increased the odds by 76% (aOR, 1.76; 95% CrI, 1.60–1.93) and more than twofold (aOR, 2.26; 95% CrI, 2.02–2.52), respectively (Supplementary Figure 7).

## Discussion

This study provides a comprehensive evaluation of four widely available general-purpose multimodal LLMs across 153 histopathology cases spanning 20 organs/systems, extending assessment beyond diagnostic accuracy to the nature, burden, and clinical consequences of model failures. Model behavior varied substantially across LLMs and case characteristics, and failures were common not only when diagnoses were wrong but also when the final diagnosis was correct. Misinterpretation and fabricated histologic features were the dominant failure modes, while higher error and hallucination burdens were strongly associated with diagnostic inaccuracy and poorer overall performance. Clinical impact also varied across models and case types, and multivariable analysis showed that diagnostic correctness was the strongest determinant of high clinical impact, with hallucination burden contributing independently to risk. Diagnostic accuracy alone is therefore insufficient to characterize the safety and reliability of LLMs in histopathology, and evaluation frameworks should also consider pathology-specific failure modes, their cumulative burden, and their potential clinical consequences.

Against this broader safety context, diagnostic accuracy in our study was modest overall, reaching 48.9% for strictly correct diagnoses and 60.1% when partially correct diagnoses were included. These values are somewhat lower than those reported in several organ-specific evaluations. For example, Mazzucchelli et al. reported 88% diagnostic accuracy for ChatGPT-4o in glioma image interpretation^25^, Jiang et al. found 79.6% accuracy for ChatGPT-4o across 250 lung tumor images^26^, and Alshammari et al. reported accuracies exceeding 90% for three LLMs in the assessment of oral lesions^27^. However, other studies have demonstrated substantially lower performance, particularly when cases are more challenging. Nguyen et al. reported accuracies of 18.7% for GPT-4o and 31.6% for o3 in oral and maxillofacial pathology cases, while an experienced pathologist achieved only 28.3% on the same image set, underscoring the intrinsic difficulty of the cases^28^. Performance also appears to decline when broader and more heterogeneous pathology domains are considered. In a multi-organ study spanning 14 organs, Apornvirat et al. reported an accuracy of 48.6% for ChatGPT-4 in microscopic description and diagnosis, closely resembling the overall performance observed here^16^. Within our own cohort, accuracy likewise varied markedly between models, with LLM4 performing best (58.2%) and LLM3 worst (36.6%). The wide range of reported performance across studies therefore likely reflects not only differences between models, but also variation in organ system, disease spectrum, case complexity, and task design.

This heterogeneity was also evident across case characteristics within our dataset. Diagnostic performance was higher in neoplastic diseases and in cases with diagnostically sufficient images, suggesting that general-purpose LLMs may be better suited to entities with distinctive and well-established morphologic patterns. Tumors such as clear cell renal cell carcinoma of the kidney, basal cell carcinoma of the skin, papillary thyroid carcinoma, invasive urothelial carcinoma, and squamous cell carcinoma of the uterine cervix, all of which were accurately diagnosed by all four LLMs in our study, often display characteristic architectural or cytologic features that may provide stronger visual cues for association with familiar diagnostic labels. By contrast, performance declined in non-neoplastic and diagnostically more complex cases, particularly when additional images or ancillary studies were required. Such cases frequently involved infectious, inflammatory, or reactive, or processes in which diagnosis depends not only on pattern recognition but also on lesion distribution, subtle diagnostic thresholds, clinical context, or exclusion of close mimics. Importantly, cases classified as requiring additional diagnostic support are intrinsically difficult even for practicing pathologists, because a confident diagnosis cannot usually be reached from the available images alone. The lower LLM performance in this setting therefore likely reflects, at least in part, the inherent incompleteness of the available evidence rather than a limitation unique to the models.

However, whether a model ultimately reached the correct diagnosis captured only part of its performance. Waqas et al. similarly showed that, despite broadly comparable factual accuracy across LLMs, substantial differences could appear in coherence, analytical depth, and pathology-specific reasoning strategies^24^. Our findings extend this concept to image-grounded diagnostic outputs: 66.2% of strictly correct diagnoses still contained at least one error and 59.9% contained a hallucination. These phenomena were even more prevalent among incorrect diagnoses, where 97.5% contained an error and 95.1% a hallucination. Thus, errors and hallucinations were strongly associated with diagnostic failure but were by no means restricted to incorrect answers. A correct final diagnosis cannot therefore be assumed to imply that the accompanying microscopic description or explanatory content is reliable; conversely, a reliable description does not necessarily guarantee a correct final diagnosis, as demonstrated in a study evaluating LLM performance in thyroid cytology specimens.^29^

The nature of these accompanying failures helps explain why diagnostic correctness alone may be misleading. Misinterpretation, omission, and fabricated features accounted for most abnormalities observed across LLM outputs. Misinterpretations frequently involved assigning an incorrect identity or significance to genuine histologic findings. These ranged from confusion between cell types and tissue components, such as interpreting Paget cells as squamous cells or adipocytes in angiomyolipoma as vacuolated spindle cells, to incorrect characterization of architecture or extracellular material, such as describing cords of invasive lobular carcinoma as a storiform pattern or mucin as hyalinized stroma. Artifacts and pathologic findings were also occasionally confused, including retraction artifact being interpreted as lymphovascular invasion and blood as keratinization. Similar feature-level limitations have been reported in oral histopathology, where agreement between multimodal LLMs and expert pathologists was particularly poor for some architectural and invasion-related features^30^. Omissions showed a complementary pattern, with models failing to report diagnostically relevant findings such as tumor differentiation, necrosis, giant cells, architectural features, or clinically relevant indicators of tumor extent including tissue invasion, lymphovascular invasion, and perineural invasion. Fabricated features were more heterogeneous and particularly concerning because they often appeared to provide additional morphologic support for the model’s favored diagnosis despite not being present in the images. In some cases, this appeared to be reinforced by anchoring to the clinical information, a phenomenon similarly reported in multimodal medical imaging models, where provision of clinical context can increase reliance on textual cues and contribute to false imaging findings^31^. In our study, for example, a pheochromocytoma in a patient with a history of lung cancer was interpreted by all models as metastatic lung adenocarcinoma, accompanied by hallucinated features supporting that diagnosis. Similarly, a papillary renal cell carcinoma clinically suspected to be a tumor arising from the renal pelvis was diagnosed by all models as invasive urothelial carcinoma, with the outputs describing invasion of renal pelvic tissue that was not demonstrated in the provided images. These patterns suggest that LLM failures in histopathology are not limited to isolated factual mistakes, but may arise from incorrect interpretation of real morphology, failure to recognize relevant findings, and generation of diagnosis-concordant features shaped by an initially favored diagnostic hypothesis or clinical context.

These feature-level failures also varied according to the amount of diagnostic information available. Hallucination rates increased as cases required greater diagnostic support, suggesting that limited or ambiguous evidence may have encouraged models to fill informational gaps with unsupported features or inferences. Previous studies have similarly shown that insufficient diagnostic evidence increases uncertainty in LLM-based diagnosis, while greater model uncertainty has been associated with hallucination-prone responses in medical image interpretation^32,33^. By contrast, error rates remained consistently high regardless of whether the available material was considered sufficient for diagnosis, indicating that misinterpretation and omission were not confined to difficult or information-limited cases. This difference suggests that hallucinations may be more sensitive to uncertainty or missing diagnostic information, whereas interpretation errors can occur even when adequate morphologic evidence is present. However, these associations are descriptive and should not be interpreted as demonstrating a causal effect of diagnostic insufficiency on hallucination generation. From an implementation perspective, these observations argue against forcing a definitive LLM diagnosis when the available material is incomplete or diagnostically insufficient. General-purpose multimodal LLMs would be most appropriately used as pathologist’s decision-support tools receiving histologic images together with relevant clinical context, with a trained pathologist responsible for assessing image adequacy, validating generated morphologic findings, and accepting or rejecting the proposed diagnosis. When image quality or diagnostic information is inadequate, expressing uncertainty or recommending additional sampling or ancillary studies would be preferable to generating unsupported diagnostic conclusions.

Beyond case difficulty, model-specific response behavior further shaped the safety profile of the outputs. LLM4, despite relatively high diagnostic accuracy, had higher adjusted odds than LLM3 of producing an erroneous or hallucinative output while remaining diagnostically correct; this may reflect its tendency to generate lengthy microscopic descriptions, thereby creating more opportunities to introduce unsupported features. Conversely, LLM2 produced the shortest outputs overall and had the lowest error rate and error burdens and was among the models with the lowest hallucination rates and burdens, suggesting that greater conciseness may reduce opportunities for failure. This was reflected in the safety classifications: LLM2 had the highest proportion of safe-correct outputs, whereas LLM3 had the lowest and also the highest proportion of dangerous-wrong outputs. A safe-correct output did not necessarily imply complete absence of minor errors or hallucinations, but rather a strictly correct diagnosis without high burden errors or hallucinations and with a clinical impact score of ≤2. LLM3 consistently showed the least favorable profile across multiple dimensions, including the lowest diagnostic accuracy and overall score, higher odds of an incorrect diagnosis, the greatest error and hallucination burden, and the highest clinical impact. The clinical consequences of these failures were particularly evident in non-neoplastic cases, where some lesions were misclassified as neoplastic and therefore received higher impact scores because benign-to-malignant errors were considered more consequential. Diagnostic accuracy alone therefore provides an incomplete measure of model safety, as response length, error burden, hallucination propensity, and the clinical consequences of specific failures all contribute to the overall risk profile of LLM-generated pathology interpretations.

These model-level differences were mirrored in the multivariable analysis of clinical impact. Diagnostic correctness was the strongest determinant, with partially correct and incorrect outputs showing substantially higher odds of high impact consequences. This association is expected because diagnostic accuracy was inherently considered during clinical impact scoring: an incorrect or incomplete diagnosis, particularly when it could alter management, was more likely to receive a higher impact score. More noteworthy was the independent association of sporadic and rare diseases with greater clinical impact. In these less common entities, less familiar or less stereotypical morphologic patterns may have increased the tendency of LLMs to generate unsupported features or inferences, which in turn contributed to diagnostic errors and amplified their potential clinical consequences. This pattern is consistent with a reinforcing sequence in which diagnostic difficulty promotes unsupported generation, that generation further distorts interpretation, and the resulting diagnostic error increases clinical impact. Although the present analysis cannot establish such a sequence causally, disease rarity appears to represent an important potential vulnerability when multimodal LLMs are applied to diagnostic pathology.

This study had several limitations. First, although all scoring pathologists received detailed and standardized instructions, assignment of some errors and hallucinations to specific subtypes may have involved subjective judgment and may not have been entirely uniform across reviewers. Nevertheless, the study primarily captures the occurrence of objectively incorrect, unsupported, or missing information, and uncertainty in subtype classification does not negate the presence of the underlying failure. Second, the dataset was intentionally assembled to encompass a broad spectrum of diagnoses rather than as a consecutive, prevalence-based clinical series, and case numbers were uneven across institutions, organs/systems, and disease categories. Performance estimates therefore characterize the selected diagnostic spectrum and should not be interpreted as prevalence-weighted estimates of real-world diagnostic performance; the relatively small number of cases in some organs/systems also increases statistical uncertainty in organ-specific comparisons. The study was not designed to evaluate demographic subgroup fairness, and potential differences in model performance across age, sex, or other patient characteristics were not assessed. In addition, representative screenshots were selected by one pathologist from whole slide images. Third, the models were evaluated using selected histologic images rather than complete whole-slide examination and the full spectrum of ancillary data available in routine practice; performance therefore reflects interpretation under the information provided and may differ in real-world diagnostic workflows. Finally, the study represents a snapshot of rapidly evolving commercial models, and subsequent model updates, changes in multimodal capabilities, or different prompting strategies could alter performance. Future studies should validate these findings in larger, more balanced multi-institutional cohorts incorporating diverse scanning platforms and whole-slide inputs, as well as repeated inference to assess reproducibility.

In conclusion, general-purpose multimodal LLMs can achieve meaningful diagnostic performance in histopathology, but their outputs remain vulnerable to frequent errors and hallucinations with variable clinical impact. These failures differed substantially across models and could persist even when the final diagnosis was correct, highlighting the limitations of diagnostic accuracy as a standalone measure of safety. Comprehensive evaluation of error type, burden, and clinical consequence is therefore essential before these models can be reliably integrated into pathology practice, where they should remain assistive tools under pathologist oversight.

## Methods

### Study design and ethics

This retrospective multicenter study evaluated the diagnostic performance of general-purpose multimodal LLMs in histopathology, and assessed pathology-relevant errors and hallucinations, their burden, and their potential clinical impact. These outcomes were prespecified human-evaluated measures assessed against a pathology reference standard. The study was reported in accordance with the TRIPOD-LLM reporting guidelines^34^.

The study was approved by the Ethics Committee of Nagasaki University (approval no. 26052204), with approval covering both participating institutions, and the requirement for patient informed consent was waived. The study was not prospectively registered. Patients and the public were not involved in the study design, conduct, analysis, or reporting. An internal study protocol was prepared by the coordinating investigator but was not prospectively published or deposited in a public repository. Participating pathologists received detailed written instructions for case evaluation and scoring rather than the full study protocol.

### Case selection and dataset preparation

The evaluation dataset comprised retrospective histopathology cases from Nagasaki University Hospital, Nagasaki, Japan, and Kameda Medical Center, Kamogawa, Japan, collected between March 2020 and January 2026. Cases were selected based on the primary diagnosis documented in the pathology report, with the aim to include a broad range of neoplastic and non-neoplastic conditions, while ensuring that no consensus diagnosis was represented more than once within the same organ system.. Cases with a non-definitive or uncertain diagnosis or without an available high-quality WSI were excluded. The initial dataset contained 158 cases; five cases (one endometrial, one lung, two skin, and one soft-tissue case) were subsequently excluded because a consensus reference diagnosis could not be established, resulting in 153 cases spanning 20 organs/systems for final analysis. Dataset characteristics are summarized in Table 1.

**Table 1.** Characteristics of the evaluation dataset.

| **Characteristic** | **Category** | **No. of cases** |
| --- | --- | --- |
| **Total cases** |  | **153** |
| **Institution** | Kameda Medical Center | 141 |
|  | Nagasaki University Hospital | 12 |
| **Sex** | Female | 83 |
|  | Male | 70 |
| **Age, years** | Mean | 58.27 |
|  | Range | 2–86 |
| **Disease category** | Neoplastic | 122 |
|  | Non-neoplastic | 31 |
| **Disease rarity** | Common | 67 |
|  | Sporadic | 63 |
|  | Rare | 23 |
| **Diagnostic efficiency** | Fully sufficient | 55 |
|  | Additional support desirable | 40 |
|  | Additional support required | 58 |
| **Organs/systems represented** |  | **20** |

All retained cases constituted the evaluation dataset; no study-specific training, fine-tuning, model development, or development/validation/test split was used. The observed distributions of disease category, disease rarity, and diagnostic-efficiency groups were retained without over-sampling, under-sampling, or other resampling procedures. No data were missing for variables included in the analyses, and no imputation was performed.

### Histologic image and clinical data preparation

All cases had been digitized before the original diagnosis using a Philips UltraFast Scanner (Philips Digital Pathology Solution, Amsterdam, the Netherlands). For each case, representative screenshots were obtained from one or two hematoxylin and eosin-stained WSIs at magnifications ranging from ×1 to ×40 (×1, ×2, ×5, ×10, ×20, and ×40). Each case included 6–17 screenshots, with more images used for morphologically heterogeneous cases to capture the relevant histologic features across magnifications. All screenshots were 1906 × 879 pixels, had a resolution of 96 dpi, and were saved in PNG format. No additional image enhancement, resizing, or preprocessing was performed.

Case and screenshot selection was performed by one pathologist (KL), with the aim of capturing representative histologic features across multiple magnifications. Images were coded and organized into one folder per case and stored on Google Drive (Google LLC, Mountain View, CA, USA).

Age, sex, the clinical diagnosis provided by the treating clinician, and a brief clinical history leading to pathological examination were retrieved from the laboratory information system. All direct identifiers, including patient and hospital names, were removed before review. Clinical information and links to the corresponding image folders were compiled in Google Sheets (Google LLC).

### LLMs and inference procedure

Four general-purpose multimodal LLMs were evaluated: ChatGPT-5.3 (OpenAI, San Francisco, CA, USA; LLM1), Gemini 3 (Google DeepMind, London, UK; LLM2), Grok 4.20 (SpaceXAI, Palo Alto, CA, USA; LLM3), and Claude Opus 4.6 (Anthropic, San Francisco, CA, USA; LLM4). The models’ training cutoff dates, dates of most recent training, and detailed proprietary information regarding model architectures, model-building procedures, and alignment strategies were not available to the investigators, and no such model-level procedures were modified as part of this study.

For each case, all four LLMs received the same screenshots in the same order, and cases themselves were presented in the same sequence across models. Each LLM was instructed to generate a microscopic description and provide the single most probable diagnosis. The prompt was developed by one pathologist (KL) after preliminary testing of alternative prompts that did not consistently elicit the required output. No evaluation cases were used during prompt optimization, and the final prompt was fixed before evaluation:

> *You are a pathologist. This is a series of pathological images at different magnifications of [name of specimen, sampling method] from a [age, gender] patient. The patient presents with [clinical information provided]. The clinical diagnosis is [clinical diagnosis provided]. Make a list of all microscopic features present in those images, and provide the most fitting single diagnosis based on the microscopic features you found.*

LLM inference was performed between February and March 2026 through the models’ respective proprietary web interfaces. Model-specific inference parameters, including temperature, maximum token length, and random seed, were not modified from the default interface settings. Only a single inference was evaluated for each case, and cases within an organ/system were processed within the same chat session with model memory enabled; consequently, run-to-run variability and potential contextual carry-over between cases were not assessed. Web browsing and other external tools were disabled.

A single response was generated and retained for each case (Supplementary Figure 8). Regeneration was permitted only when the initial response failed to provide the requested microscopic description and diagnosis and instead returned a clarification question. LLM outputs were copied verbatim without editing. Across the 153 cases and four models, 612 outputs were generated.

Because inference was performed through proprietary web interfaces, information regarding the underlying computational hardware, accelerator type and number, floating-point operations, and related compute characteristics was unavailable. No local computational resources were used for LLM inference, and inference time and per-query computational cost were not systematically recorded.

### Pathologist review and reference diagnosis

Twenty-one pathologists reviewed the 153 cases, with each pathologist assigned cases from one to three organs/systems according to subspecialty expertise and experience. Reviewers had 8–30 years of diagnostic pathology experience. Each case was independently reviewed by two pathologists.

Before viewing the LLM outputs, reviewers assessed the same photomicrographs provided to the models and independently recorded a brief microscopic description and their most probable diagnosis. For cases in which the two pathologists provided discordant diagnoses, a consensus discussion was conducted to establish the reference diagnosis. Cases for which consensus could not be achieved were excluded from further analysis. The resulting consensus diagnosis served as the pathology reference standard. Pathologists also classified the diagnostic efficiency of the provided material according to whether the images were fully sufficient for a confident diagnosis, whether additional images or ancillary tests were desirable, or whether additional images or ancillary tests were required. Disease rarity was independently categorized by expert judgment as common, sporadic, or rare according to the frequency with which each diagnosis is encountered in routine pathology practice; no formal epidemiologic incidence thresholds were applied.

After completing these assessments, pathologists were allowed to review the LLM-generated microscopic descriptions and diagnoses. Reviewers were blinded to LLM identity and received detailed written instructions for the evaluation procedure; some reviewers additionally received a live videoconference explanation of the scoring process.

### Assessment of errors and hallucinations

After reviewing each LLM output, pathologists recorded the presence of predefined pathology-relevant errors (omission, misinterpretation, and internal inconsistency) and hallucinations (fabricated feature, unsupported inference, and instruction hallucination).

An omission was defined as failure to report an important microscopic feature that was clearly present and relevant to the diagnosis, including failure to identify an important tumor subtype. Misinterpretation referred to recognition of a real histologic feature but assignment of an incorrect meaning to that feature. Internal inconsistency was defined as a contradiction within the LLM’s own response.

A fabricated feature was defined as a microscopic feature described by the LLM that was not present in the provided images. Unsupported inference referred to a diagnostic or interpretive statement that could not be supported by the provided histologic images or clinical information and implicitly depended on unavailable information, such as immunohistochemical, molecular, radiologic, or laboratory findings. Instruction hallucination was defined as failure to follow the requested task accompanied by the introduction of extraneous or invented context.

Each error and hallucination subtype was scored from 0 to 3 according to the number of occurrences within an output: 0 indicated none, 1 indicated one occurrence, 2 indicated two occurrences, and 3 indicated three or more occurrences. These subtype scores were subsequently used to derive total error and hallucination burden.

### Clinical impact and overall performance assessment

Pathologists also evaluated the potential clinical consequences of each LLM output using a five-level clinical impact scale ranging from 0 to 4, with higher scores indicating greater potential harm if the output were adopted by a pathologist or clinician. This metric was intended to provide a clinically relevant assessment of output safety rather than relying on automated text-similarity measures. Primary quality and safety outcomes were directly assessed by pathologists; therefore, correlation with a separate automated text-quality metric was not applicable.

Clinical impact was scored as follows: 0 (none), no meaningful clinical impact, such as a stylistic issue or minor wording problem; 1 (minor), a trivial error not expected to alter management; 2 (moderate), an error that could lead to additional testing without directly changing treatment, such as an unnecessary immunohistochemistry panel; 3 (major), an error with the potential to alter treatment or surgical management, such as misclassification of benign versus malignant disease or an incorrect grade affecting therapy; and 4 (critical), an error with potential for life-threatening harm, such as failure to recognize invasion or incorrect classification of metastatic disease leading to undertreatment.

Reviewers additionally assigned an overall LLM performance score from 1 (poor) to 5 (excellent), integrating the quality of the microscopic description and final diagnosis.

### Consensus and diagnostic correctness

LLM diagnostic correctness was classified by one pathologist (KL) using the consensus diagnosis as the reference standard. A response was considered correct when the final LLM diagnosis matched the consensus diagnosis, partially correct when it captured the principal diagnosis but was incomplete or contained an incorrect secondary qualification, such as a missing subtype or a correct diagnosis followed by an incorrect subtype or assessment, and incorrect when the final diagnosis differed substantially from the consensus diagnosis.

The higher score was retained for diagnostic efficiency, disease rarity, error and hallucination subtype scores, and clinical impact. For overall LLM performance, the mean of the two reviewers’ scores was used for analysis.

### Statistical analysis

Statistical analyses were performed primarily at the LLM output level. Each of the 153 cases generated four correlated outputs, one from each LLM. Categorical variables are reported as counts and percentages, and numerical or ordinal variables as mean ± standard deviation (SD) with 95% confidence intervals (CIs), as appropriate. All frequentist tests were two-sided with a significance threshold of *p* < 0.05. Holm adjustment was applied to multiple pairwise comparisons.

Strict diagnostic accuracy was defined as the proportion of outputs classified as correct, whereas partial diagnostic accuracy included both correct and partially correct diagnoses. Because all four LLMs evaluated the same cases, differences in binary outcomes among models, including diagnostic accuracy and the presence of individual error or hallucination types, were assessed using Cochran’s Q test followed, when significant, by Holm-adjusted pairwise McNemar tests. Comparisons across independent case characteristics, including disease category, disease rarity, and diagnostic efficiency, were performed using Fisher’s exact or omnibus contingency-table tests, as appropriate.

An error or hallucination was considered present when any corresponding subtype score was ≥1. Total error burden was calculated by summing omission, misinterpretation, and internal-inconsistency scores, and total hallucination burden by summing fabricated-feature, unsupported-inference, and instruction-hallucination scores, yielding burden scores from 0 to 9. High burden was defined as a total error or hallucination burden score of 3 or greater. Differences in burden, clinical impact, and overall LLM score among the four paired models were assessed using Friedman tests followed by Holm-adjusted Wilcoxon signed-rank tests. Comparisons across independent case groups were performed using Mann–Whitney U or Kruskal–Wallis tests, as appropriate. Generalized estimating equation (GEE) models were used for pooled analyses requiring adjustment for the four correlated outputs per case. Gaussian GEE models with an identity link evaluated error and hallucination burden across case characteristics and diagnostic correctness, while logistic GEE models were used for binary outcomes. Partially correct diagnoses were excluded from comparisons restricted to strictly correct versus incorrect outputs.

The relationship between cumulative burden and diagnostic performance was additionally examined using proportional-odds ordinal logistic regression for each LLM, with diagnostic correctness modeled as incorrect, partially correct, or correct and burden entered as a continuous predictor. Associations between individual error or hallucination subtypes and clinical impact were assessed using simultaneous case-clustered GEE models adjusted for LLM identity. Mean clinical impact was modeled using Gaussian GEE, whereas high clinical impact, defined as a score of 3–4, was modeled using logistic GEE. Clinically relevant output patterns were also summarized by classifying outputs as safe correct or dangerous wrong according to diagnostic correctness, clinical impact, and error/hallucination burden, with differences among LLMs evaluated using paired binary-outcome comparisons.

Multivariable analyses were performed for five prespecified adverse outcomes: incorrect diagnosis, any error, any hallucination, high clinical impact, and low overall LLM performance (score ≤2). Bayesian mixed-effects logistic regression models included random intercepts for case and organ/system to account for within-case correlation and between-organ/system heterogeneity. Models for incorrect diagnosis, error, and hallucination included LLM identity, disease category, disease rarity, and diagnostic efficiency as fixed effects; models for high clinical impact and low overall performance additionally included diagnostic correctness and total error and hallucination burden. Bayesian models were fitted using BinomialBayesMixedGLM with weakly informative priors and variational Bayes estimation. Results are reported as adjusted odds ratios (aORs) with 95% credible intervals (CrIs), with associations for non-LLM predictors considered supported when the CrI excluded 1. The overall adjusted effect of LLM identity was evaluated using an approximate three-degree-of-freedom joint Wald test, followed by all six pairwise LLM contrasts with Holm-adjusted two-sided *p* values.

Analyses were performed using Python version 3.13.5, with pandas 2.2.3 and NumPy 2.3.5 for data management, statsmodels 0.14.6 for regression and GEE analyses, and SciPy 1.17.0 for additional statistical calculations.

## Author contributions

KL and JF conceived the study. KL designed the study, and KL, EU, AB, and JF critically revised the study design. KL and JF collected all cases, performed data curation, and prompted LLMs. KL, SA, AA, SB, AH, HK, JK, MK, TL, SM, JM, HP, DGP, AS, SS, KS, MS, YT, IT, QW, and JF reviewed the pathological images, provided microscopic descriptions and diagnoses, and evaluated the LLM outputs. KL analyzed and interpreted the data, performed the statistical analyses and data visualization, and drafted the manuscript. All authors critically revised the manuscript, read and approved the final version.

## Acknowledgements

This study was funded by the AI Industry Strategy Office, IT Industry Division, Commerce and Information Policy Bureau, Ministry of Economy, Trade and Industry. The funder played no role in study design, data collection, analysis and interpretation of data, or the writing of this manuscript.

## Data availability

The datasets analyzed during the current study are not publicly available due to institutional restrictions on sharing clinical and histopathological data but are available from the corresponding author on reasonable request and subject to applicable institutional and ethical approvals.

## Code availability

The analysis code used to generate the statistical results reported in this study is publicly available at https://github.com/krislami/llm-pathology-errors-hallucinations. The repository includes code for data preprocessing, derivation of study outcomes, diagnostic-performance analyses, error and hallucination analyses, burden and clinical-impact analyses, generalized estimating equation models, proportional-odds models, Bayesian mixed-effects logistic regression, multiple-comparison adjustment, and generation of analysis outputs and figures.

## Supplementary Figure Legends

**Supplementary Figure 1. Representative examples of pathology-relevant errors and hallucinations in LLM outputs.**

Representative histopathological examples illustrating the six predefined failure subtypes. Top row, errors: omission, illustrated by failure to report atypical mitoses and giant cells; misinterpretation, illustrated by epidermis being interpreted as ductal structures; and internal inconsistency, illustrated by an output stating the absence of high-grade features but subsequently providing a malignant diagnosis. Bottom row, hallucinations: fabricated feature, illustrated by reporting squamous differentiation that was not present; unsupported inference, illustrated by diagnosing neuroendocrine carcinoma without supporting evidence; and instruction hallucination, illustrated by discussion of treatment effects despite these not being requested. LLM, large language model.

**Supplementary Figure 2. Error and hallucination rates according to disease category.**

Overall prevalence of errors and hallucinations and model-specific error and hallucination rates are shown for neoplastic and non-neoplastic cases. Error bars represent 95% Wilson binomial confidence intervals. LLM, large language model.

**Supplementary Figure 3. Error and hallucination rates according to disease rarity.**

Overall prevalence of errors and hallucinations and model-specific error and hallucination rates are shown for common, sporadic, and rare diseases. Error bars represent 95% Wilson binomial confidence intervals. Er, error; Ha, hallucination; LLM, large language model.

**Supplementary Figure 4. Error and hallucination rates according to diagnostic efficiency.**

Overall prevalence of errors and hallucinations and model-specific error and hallucination rates are shown according to the diagnostic efficiency of the provided material. Error bars represent 95% Wilson binomial confidence intervals. Er, error; Ha, hallucination; LLM, large language model.

**Supplementary Figure 5. Clinical impact score by organ.**

Mean clinical impact scores are shown for each organ across all LLM outputs. Points represent mean clinical impact scores and error bars represent 95% confidence intervals. Clinical impact was scored from 0 to 4, with higher scores indicating greater potential clinical consequences. CNS, central nervous system; LLM, large language model.

**Supplementary Figure 6. Overall LLM performance scores across models and case characteristics.**

a, Mean overall performance score for each LLM. b, Mean overall scores according to disease category (neoplastic and non-neoplastic). c, Mean overall scores according to disease rarity (common, sporadic, and rare). d, Mean overall scores according to diagnostic efficiency of the provided material (fully sufficient, additional support desirable, and additional support required). e, Mean overall scores for each LLM across the 20 organs. Overall performance was scored from 1 (poor) to 5 (excellent), with higher scores indicating better performance. Points represent mean scores and error bars represent 95% confidence intervals. Brackets indicate statistically significant pairwise comparisons; one asterisk indicates *p* < 0.05, two asterisks *p* < 0.01, and three asterisks *p* < 0.001. Only statistically significant comparisons are shown. CNS, central nervous system; LLM, large language model.

**Supplementary Figure 7. Multivariable analysis of factors associated with a low overall LLM score.**

Forest plot showing adjusted associations with a low overall LLM performance score, defined as a score of 2 or lower. Blue points represent adjusted odds ratios (aORs) and horizontal bars represent 95% credible intervals (CrIs); the vertical dashed line indicates an aOR of 1. LLM effects are presented as all pairwise contrasts, with Holm-adjusted *P* values; the overall adjusted LLM effect is shown above the plot. Reference categories for the remaining predictors were neoplastic disease, common disease, fully sufficient diagnostic material, and correct diagnosis. Error and hallucination burdens were modeled per one-point increase. Values greater than 1 indicate higher adjusted odds of a low LLM score, whereas values below 1 indicate lower odds. Bold estimates denote statistically supported associations. Models were estimated using mixed-effects logistic regression with random intercepts for case and organ. LLM, large language model; aOR, adjusted odds ratio; CrI, credible interval.

**Supplementary Figure 8. Representative LLM outputs for the same breast biopsy case.** Representative verbatim outputs generated from the same histopathological images and clinical information. a, LLM2 generated a correct diagnosis of invasive lobular carcinoma, with one omission identified in the accompanying microscopic description. b, LLM3 generated an incorrect diagnosis of spindle cell metaplastic carcinoma and the output contained three omissions, three misinterpretations, and three fabricated features. These examples illustrate how model outputs could differ substantially in both diagnostic correctness and the number and type of pathology-relevant failures despite receiving identical input images and clinical context. LLM, large language model.

