## Supplementary Table 1 for "Errors, Hallucinations, and Clinical Impact of General-Purpose Multimodal Large Language Models in Histopathology"

**Supplementary Table 1. Evaluation cases and consensus diagnoses**

| **Organ** | **Consensus diagnosis** | **Disease category** | **Disease rarity** |
| --- | --- | --- | --- |
| Adrenal gland | Adrenal cortical adenoma | Neoplastic | Sporadic |
| Adrenal gland | Pheochromocytoma | Neoplastic | Sporadic |
| Breast | Paget disease | Neoplastic | Common |
| Breast | Invasive lobular carcinoma | Neoplastic | Common |
| Breast | Metaplastic carcinoma | Neoplastic | Sporadic |
| Breast | Solid papillary carcinoma | Neoplastic | Sporadic |
| Breast | Invasive breast carcinoma of no special type | Neoplastic | Common |
| Breast | Invasive breast carcinoma of no special type with medullary features | Neoplastic | Sporadic |
| Breast | Ductal carcinoma in situ | Neoplastic | Sporadic |
| Breast | Benign phyllodes tumor | Neoplastic | Sporadic |
| Breast | Nipple adenoma | Neoplastic | Rare |
| Central nervous system | Fibrous meningioma | Neoplastic | Common |
| Central nervous system | Secretory meningioma | Neoplastic | Common |
| Central nervous system | Glioblastoma, WHO grade 4 | Neoplastic | Common |
| Central nervous system | Astrocytoma | Neoplastic | Common |
| Central nervous system | Psammomatous meningioma | Neoplastic | Common |
| Central nervous system | Pituitary adenoma | Neoplastic | Common |
| Central nervous system | Schwannoma | Neoplastic | Common |
| Central nervous system | Atypical meningioma | Neoplastic | Sporadic |
| Central nervous system | Meningothelial meningioma | Neoplastic | Common |
| Colon | Cytomegalovirus colitis | Non-neoplastic | Sporadic |
| Colon | Tubular adenocarcinoma | Neoplastic | Common |
| Colon | Ulcerative colitis | Non-neoplastic | Sporadic |
| Colon | Intestinal spirochetosis | Non-neoplastic | Sporadic |
| Colon | Medullary carcinoma | Neoplastic | Rare |
| Colon | Traditional serrated adenoma | Neoplastic | Sporadic |
| Colon | Neuroendocrine tumor, grade 1 | Neoplastic | Sporadic |
| Colon | Sessile serrated lesion | Neoplastic | Common |
| Colon | Leiomyosarcoma | Neoplastic | Rare |
| Colon | Low-grade tubular adenoma | Neoplastic | Common |
| Colon | Amebic colitis | Non-neoplastic | Rare |
| Endometrium | Stromal breakdown | Non-neoplastic | Common |
| Endometrium | Adenomyosis | Non-neoplastic | Common |
| Endometrium | Serous carcinoma | Neoplastic | Sporadic |
| Endometrium | Products of conception | Non-neoplastic | Common |
| Endometrium | Simple hyperplasia | Neoplastic | Common |
| Endometrium | Leiomyosarcoma | Neoplastic | Common |
| Endometrium | Endometrioid carcinoma | Neoplastic | Common |
| Endometrium | Carcinosarcoma | Neoplastic | Sporadic |
| Endometrium | Endometrial stromal sarcoma | Neoplastic | Rare |
| Esophagus | Squamous dysplasia | Neoplastic | Common |
| Esophagus | Squamous papilloma | Neoplastic | Common |
| Esophagus | Esophageal candidiasis | Non-neoplastic | Rare |
| Esophagus | Squamous cell carcinoma | Neoplastic | Common |
| Esophagus | Eosinophilic esophagitis | Non-neoplastic | Sporadic |
| Esophagus | Granular cell tumor | Neoplastic | Sporadic |
| Esophagus | Carcinosarcoma | Neoplastic | Rare |
| Esophagus | Herpes esophagitis | Non-neoplastic | Sporadic |
| Esophagus | Barrett esophagus | Non-neoplastic | Common |
| Esophagus | Glycogenic acanthosis | Non-neoplastic | Common |
| Kidney | Clear cell renal cell carcinoma | Neoplastic | Common |
| Kidney | Oncocytoma | Neoplastic | Sporadic |
| Kidney | Angiomyolipoma | Neoplastic | Sporadic |
| Kidney | Papillary neoplasm with reverse polarity | Neoplastic | Rare |
| Kidney | Chromophobe renal cell carcinoma | Neoplastic | Sporadic |
| Kidney | Mucinous tubular and spindle cell carcinoma | Neoplastic | Rare |
| Kidney | Nephroblastoma | Neoplastic | Rare |
| Kidney | Papillary renal cell carcinoma | Neoplastic | Common |
| Liver | Intrahepatic cholangiocarcinoma | Neoplastic | Sporadic |
| Liver | Cavernous hemangioma | Neoplastic | Sporadic |
| Liver | Steatohepatitis | Non-neoplastic | Sporadic |
| Liver | Hepatocellular carcinoma | Neoplastic | Sporadic |
| Liver | Cirrhosis | Non-neoplastic | Sporadic |
| Lung | Micropapillary adenocarcinoma | Neoplastic | Sporadic |
| Lung | Small cell lung carcinoma | Neoplastic | Common |
| Lung | Acute lung injury | Non-neoplastic | Common |
| Lung | Necrotizing granulomatous inflammation | Non-neoplastic | Common |
| Lung | Squamous cell carcinoma | Neoplastic | Common |
| Lung | Pleuroparenchymal fibroelastosis | Non-neoplastic | Rare |
| Lung | Hamartoma | Neoplastic | Common |
| Lung | Usual interstitial pneumonia | Non-neoplastic | Common |
| Ovary | Clear cell carcinoma | Neoplastic | Sporadic |
| Ovary | Mucinous cystadenoma | Neoplastic | Common |
| Ovary | Mature cystic teratoma | Neoplastic | Common |
| Ovary | Dysgerminoma | Neoplastic | Rare |
| Ovary | High-grade serous carcinoma | Neoplastic | Common |
| Ovary | Struma ovarii | Neoplastic | Sporadic |
| Ovary | Follicular cyst | Neoplastic | Common |
| Ovary | Adult granulosa cell tumor | Neoplastic | Sporadic |
| Ovary | Mucinous borderline tumor | Neoplastic | Sporadic |
| Pancreas | Intraductal papillary mucinous neoplasm | Neoplastic | Sporadic |
| Pancreas | Ductal adenocarcinoma | Neoplastic | Common |
| Pancreas | Well-differentiated neuroendocrine tumor | Neoplastic | Sporadic |
| Pancreas | Acinar cell carcinoma | Neoplastic | Rare |
| Prostate | Benign prostatic hyperplasia | Non-neoplastic | Common |
| Prostate | Acinar adenocarcinoma, Gleason score 3+3=6 | Neoplastic | Common |
| Prostate | Basal cell hyperplasia | Non-neoplastic | Sporadic |
| Prostate | Acinar adenocarcinoma with ductal adenocarcinoma component | Neoplastic | Sporadic |
| Prostate | Granulomatous prostatitis | Non-neoplastic | Sporadic |
| Prostate | Acinar adenocarcinoma, Gleason score 4+5=9 | Neoplastic | Sporadic |
| Salivary gland | Myoepithelioma | Neoplastic | Rare |
| Salivary gland | Warthin tumor | Neoplastic | Common |
| Salivary gland | Mucoepidermoid carcinoma | Neoplastic | Sporadic |
| Salivary gland | Sialadenitis consistent with Sjögren syndrome | Non-neoplastic | Sporadic |
| Salivary gland | Adenoid cystic carcinoma | Neoplastic | Sporadic |
| Salivary gland | Pleomorphic adenoma | Neoplastic | Common |
| Salivary gland | Salivary duct carcinoma | Neoplastic | Rare |
| Skin | Seborrheic keratosis | Neoplastic | Common |
| Skin | Basal cell carcinoma, infiltrating type | Neoplastic | Common |
| Skin | Merkel cell carcinoma | Neoplastic | Rare |
| Skin | Neurofibroma | Neoplastic | Sporadic |
| Skin | Actinic keratosis | Neoplastic | Sporadic |
| Skin | Psoriasis | Non-neoplastic | Sporadic |
| Skin | Melanoma | Neoplastic | Sporadic |
| Skin | Schwannoma | Neoplastic | Sporadic |
| Skin | Malignant peripheral nerve sheath tumor | Neoplastic | Rare |
| Skin | Bullous pemphigoid | Non-neoplastic | Sporadic |
| Skin | Eccrine poroma | Neoplastic | Sporadic |
| Skin | Intradermal nevus | Neoplastic | Common |
| Soft tissue | Lipoma | Neoplastic | Common |
| Soft tissue | Angioleiomyoma | Neoplastic | Common |
| Soft tissue | Glomus tumor | Neoplastic | Common |
| Soft tissue | Osteosarcoma | Neoplastic | Common |
| Soft tissue | Tenosynovial giant cell tumor | Neoplastic | Common |
| Soft tissue | Well-differentiated liposarcoma | Neoplastic | Common |
| Soft tissue | Gout | Non-neoplastic | Common |
| Soft tissue | Dedifferentiated chondrosarcoma | Neoplastic | Common |
| Stomach | MALT lymphoma | Neoplastic | Sporadic |
| Stomach | Helicobacter pylori gastritis | Non-neoplastic | Common |
| Stomach | Amyloidosis | Non-neoplastic | Sporadic |
| Stomach | Signet ring cell carcinoma | Neoplastic | Sporadic |
| Stomach | Xanthoma | Neoplastic | Sporadic |
| Stomach | Fundic gland polyp | Neoplastic | Common |
| Stomach | Mixed adenoneuroendocrine carcinoma (MANEC) | Neoplastic | Rare |
| Stomach | Gastrointestinal stromal tumor | Neoplastic | Common |
| Stomach | Intestinal-type adenoma | Neoplastic | Common |
| Stomach | Diffuse large B-cell lymphoma | Neoplastic | Rare |
| Stomach | Chronic gastritis with intestinal metaplasia | Non-neoplastic | Sporadic |
| Testis | Embryonal carcinoma | Neoplastic | Common |
| Testis | Mixed germ cell tumor (teratoma and yolk sac tumor) | Neoplastic | Sporadic |
| Testis | Seminoma | Neoplastic | Common |
| Testis | Sertoli cell-only syndrome | Non-neoplastic | Common |
| Thyroid | Papillary thyroid carcinoma | Neoplastic | Common |
| Thyroid | Graves disease | Neoplastic | Common |
| Thyroid | Noninvasive follicular thyroid neoplasm with papillary-like nuclear features | Neoplastic | Sporadic |
| Thyroid | Medullary thyroid carcinoma | Neoplastic | Sporadic |
| Thyroid | Chronic lymphocytic thyroiditis | Non-neoplastic | Sporadic |
| Thyroid | Oncocytic carcinoma | Neoplastic | Sporadic |
| Thyroid | Follicular thyroid carcinoma | Neoplastic | Rare |
| Urinary bladder | Invasive urothelial carcinoma | Neoplastic | Sporadic |
| Urinary bladder | Inverted urothelial papilloma | Neoplastic | Sporadic |
| Urinary bladder | High-grade invasive urothelial carcinoma with squamous differentiation | Neoplastic | Sporadic |
| Urinary bladder | Cystitis cystica et glandularis | Non-neoplastic | Sporadic |
| Urinary bladder | Small cell neuroendocrine carcinoma | Neoplastic | Rare |
| Urinary bladder | Noninvasive papillary urothelial carcinoma | Neoplastic | Common |
| Uterine cervix | Endocervical polyp | Neoplastic | Common |
| Uterine cervix | High-grade squamous intraepithelial lesion | Neoplastic | Sporadic |
| Uterine cervix | HPV-associated adenocarcinoma | Neoplastic | Sporadic |
| Uterine cervix | Small cell neuroendocrine carcinoma | Neoplastic | Rare |
| Uterine cervix | Low-grade squamous intraepithelial lesion / cervical intraepithelial neoplasia 1 | Neoplastic | Common |
| Uterine cervix | Lobular endocervical glandular hyperplasia | Neoplastic | Rare |
| Uterine cervix | Keratinizing squamous cell carcinoma | Neoplastic | Common |
| Uterine cervix | Condyloma acuminatum | Neoplastic | Common |
