## Supplementary Table 2 for "Errors, Hallucinations, and Clinical Impact of General-Purpose Multimodal Large Language Models in Histopathology"

**Supplementary Table 2. Case-level patterns of diagnostic agreement among the four large language models**

| **Diagnostic agreement category** | **Organ** | **Diagnosis (LLM, where applicable)** |
| --- | --- | --- |
| **All four LLMs correct** — **29 cases (19.0%)** | Adrenal | Adrenal cortical adenoma |
|  | Breast | Invasive breast carcinoma of no special type |
|  | CNS | Pituitary adenoma; schwannoma; meningothelial meningioma |
|  | Colon | Ulcerative colitis |
|  | Esophagus | Squamous papilloma; eosinophilic esophagitis |
|  | Kidney | Clear cell renal cell carcinoma |
|  | Liver | Intrahepatic cholangiocarcinoma |
|  | Lung | Hamartoma |
|  | Ovary | Dysgerminoma; high-grade serous carcinoma |
|  | Pancreas | Intraductal papillary mucinous neoplasm |
|  | Prostate | Benign prostatic hyperplasia |
|  | Salivary gland | Sialadenitis |
|  | Skin | Basal cell carcinoma; Merkel cell carcinoma; psoriasis; bullous pemphigoid |
|  | Stomach | Signet ring cell carcinoma |
|  | Thyroid | Papillary thyroid carcinoma; Graves’ disease |
|  | Urinary bladder | Invasive urothelial carcinoma |
|  | Uterine cervix | Endocervical polyp; High-grade squamous intraepithelial lesion; HPV-associated adenocarcinoma; keratinizing squamous cell carcinoma; condyloma acuminatum |
| **Only one LLM correct** — **23 cases (15.0%)** | Breast | Paget disease (LLM2); ductal carcinoma in situ (LLM4) |
|  | Colon | Leiomyosarcoma (LLM2); amebic colitis (LLM2) |
|  | Endometrium | Serous carcinoma (LLM2); product of conception (LLM1) |
|  | Esophagus | Candida infection (LLM1); carcinosarcoma (LLM4) |
|  | Kidney | Chromophobe renal cell carcinoma (LLM2) |
|  | Ovary | Mucinous cystadenoma (LLM1); mucinous borderline tumor (LLM3) |
|  | Prostate | Acinar adenocarcinoma with ductal adenocarcinoma component (LLM1); acinar adenocarcinoma, Gleason score 4+5 (LLM4) |
|  | Salivary gland | Mucoepidermoid carcinoma (LLM2) |
|  | Skin | Actinic keratosis (LLM4) |
|  | Soft tissue | Lipoma (LLM1); angioleiomyoma (LLM3) |
|  | Stomach | Mixed adenoneuroendocrine carcinoma (MANEC) (LLM4); intestinal-type adenoma (LLM4); Diffuse large B-cell lymphoma(LLM4) |
|  | Thyroid | Medullary carcinoma (LLM2); oncocytic carcinoma (LLM4); follicular thyroid carcinoma (LLM3) |
| **No LLM strictly correct** — **22 cases (14.4%)** | Adrenal | Pheochromocytoma |
|  | CNS | Secretory meningioma |
|  | Colon | Cytomegalovirus colitis; spirochetosis; traditional serrated adenoma; tubular adenoma |
|  | Esophagus | Herpes esophagitis |
|  | Kidney | Papillary renal cell carcinoma |
|  | Lung | Usual interstitial pneumonia |
|  | Prostate | Basal cell hyperplasia; granulomatous prostatitis |
|  | Salivary gland | Myoepithelioma |
|  | Skin | Neurofibroma; eccrine poroma; intradermal nevus |
|  | Soft tissue | Gout |
|  | Stomach | Amyloidosis; xanthoma |
|  | Testis | Embryonal carcinoma |
|  | Thyroid | Chronic lymphocytic thyroiditis |
|  | Urinary bladder | Inverted urothelial papilloma |
|  | Uterine cervix | Low-grade squamous intraepithelial lesion / cervical intraepithelial neoplasia 1 |

**Notes:** For the only one LLM correct category, the LLM shown in parentheses was the only model providing the strictly correct diagnosis. The no LLM strictly correct category excludes partially correct diagnoses; thus, none of the four models provided a strictly correct diagnosis for these cases.
