## Supplementary Table 3 for "Errors, Hallucinations, and Clinical Impact of General-Purpose Multimodal Large Language Models in Histopathology"

**Supplementary Table 3. Diagnostic accuracy by organ across all four LLMs**

| **Organ** | **No. of cases** | **Strictly correct diagnosis, n/N (%)** | **Correct or partially correct diagnosis, n/N (%)** |
| --- | --- | --- | --- |
| Adrenal | 2 | 4/8 (50.0%) | 4/8 (50.0%) |
| Breast | 9 | 22/36 (61.1%) | 29/36 (80.6%) |
| CNS | 9 | 24/36 (66.7%) | 25/36 (69.4%) |
| Colon | 11 | 14/44 (31.8%) | 17/44 (38.6%) |
| Endometrium | 9 | 17/36 (47.2%) | 29/36 (80.6%) |
| Esophagus | 10 | 18/40 (45.0%) | 26/40 (65.0%) |
| Kidney | 8 | 15/32 (46.9%) | 19/32 (59.4%) |
| Liver | 5 | 9/20 (45.0%) | 15/20 (75.0%) |
| Lung | 8 | 10/32 (31.2%) | 17/32 (53.1%) |
| Ovary | 9 | 20/36 (55.6%) | 22/36 (61.1%) |
| Pancreas | 4 | 12/16 (75.0%) | 12/16 (75.0%) |
| Prostate | 6 | 8/24 (33.3%) | 11/24 (45.8%) |
| Salivary gland | 7 | 14/28 (50.0%) | 14/28 (50.0%) |
| Skin | 12 | 28/48 (58.3%) | 28/48 (58.3%) |
| Soft tissue | 8 | 13/32 (40.6%) | 17/32 (53.1%) |
| Stomach | 11 | 15/44 (34.1%) | 17/44 (38.6%) |
| Testis | 4 | 6/16 (37.5%) | 8/16 (50.0%) |
| Thyroid | 7 | 11/28 (39.3%) | 16/28 (57.1%) |
| Urinary bladder | 6 | 14/24 (58.3%) | 16/24 (66.7%) |
| Uterine cervix | 8 | 25/32 (78.1%) | 26/32 (81.2%) |

**Note:** Each case was independently evaluated by four LLMs; therefore, the denominator for each organ corresponds to four times the number of cases. CNS, central nervous system; LLM, large language model.
