## Supplementary Table 4 for "Errors, Hallucinations, and Clinical Impact of General-Purpose Multimodal Large Language Models in Histopathology"

**Supplementary Table 4. Diagnostic accuracy by organ and LLM**

| **Organ** | **No. of cases** | **LLM1, C/P/W, n (%)** | **LLM2, C/P/W, n (%)** | **LLM3, C/P/W, n (%)** | **LLM4, C/P/W, n (%)** |
| --- | --- | --- | --- | --- | --- |
| Adrenal | 2 | 1/0/1 (50.0%) | 1/0/1 (50.0%) | 1/0/1 (50.0%) | 1/0/1 (50.0%) |
| Breast | 9 | 3/3/3 (33.3%) | 6/2/1 (66.7%) | 5/2/2 (55.6%) | 8/0/1 (88.9%) |
| CNS | 9 | 6/1/2 (66.7%) | 6/0/3 (66.7%) | 5/0/4 (55.6%) | 7/0/2 (77.8%) |
| Colon | 11 | 3/2/6 (27.3%) | 5/1/5 (45.5%) | 2/0/9 (18.2%) | 4/0/7 (36.4%) |
| Endometrium | 9 | 5/3/1 (55.6%) | 4/3/2 (44.4%) | 2/3/4 (22.2%) | 6/3/0 (66.7%) |
| Esophagus | 10 | 7/2/1 (70.0%) | 3/2/5 (30.0%) | 2/2/6 (20.0%) | 6/2/2 (60.0%) |
| Kidney | 8 | 2/2/4 (25.0%) | 6/1/1 (75.0%) | 4/0/4 (50.0%) | 3/1/4 (37.5%) |
| Liver | 5 | 3/2/0 (60.0%) | 2/0/3 (40.0%) | 2/2/1 (40.0%) | 2/2/1 (40.0%) |
| Lung | 8 | 2/2/4 (25.0%) | 3/3/2 (37.5%) | 2/0/6 (25.0%) | 3/2/3 (37.5%) |
| Ovary | 9 | 5/1/3 (55.6%) | 5/0/4 (55.6%) | 4/1/4 (44.4%) | 6/0/3 (66.7%) |
| Pancreas | 4 | 4/0/0 (100.0%) | 3/0/1 (75.0%) | 1/0/3 (25.0%) | 4/0/0 (100.0%) |
| Prostate | 6 | 2/1/3 (33.3%) | 2/2/2 (33.3%) | 1/0/5 (16.7%) | 3/0/3 (50.0%) |
| Salivary gland | 7 | 4/0/3 (57.1%) | 6/0/1 (85.7%) | 2/0/5 (28.6%) | 2/0/5 (28.6%) |
| Skin | 12 | 7/0/5 (58.3%) | 7/0/5 (58.3%) | 5/0/7 (41.7%) | 9/0/3 (75.0%) |
| Soft tissue | 8 | 5/1/2 (62.5%) | 3/1/4 (37.5%) | 3/1/4 (37.5%) | 2/1/5 (25.0%) |
| Stomach | 11 | 3/1/7 (27.3%) | 3/0/8 (27.3%) | 1/1/9 (9.1%) | 8/0/3 (72.7%) |
| Testis | 4 | 0/1/3 (0.0%) | 2/0/2 (50.0%) | 2/0/2 (50.0%) | 2/1/1 (50.0%) |
| Thyroid | 7 | 2/1/4 (28.6%) | 3/2/2 (42.9%) | 3/1/3 (42.9%) | 3/1/3 (42.9%) |
| Urinary bladder | 6 | 2/1/3 (33.3%) | 5/0/1 (83.3%) | 3/1/2 (50.0%) | 4/0/2 (66.7%) |
| Uterine cervix | 8 | 6/1/1 (75.0%) | 7/0/1 (87.5%) | 6/0/2 (75.0%) | 6/0/2 (75.0%) |

**Note:** Values are presented as the number of correct/partially correct/incorrect diagnoses (C/P/W), with the percentage in parentheses representing the proportion of strictly correct diagnoses for that organ and LLM. Each LLM evaluated every case within each organ. C, correct; P, partially correct; W, incorrect; CNS, central nervous system; LLM, large language model.
