## Supplementary Table 5 for "Errors, Hallucinations, and Clinical Impact of General-Purpose Multimodal Large Language Models in Histopathology"

**Supplementary Table 5. Distribution of error and hallucination subtypes by disease category**

| **Failure subtype** | **Neoplastic**  **n = 488 outputs** | **Non-neoplastic**  **n = 124 outputs** |
| --- | --- | --- |
| **Errors** |  |  |
| Omission | 268 (54.9%) | 58 (46.8%) |
| Misinterpretation | 350 (71.7%) | 91 (73.4%) |
| Internal inconsistency | 81 (16.6%) | 24 (19.4%) |
| **Hallucinations** |  |  |
| Fabricated feature | 353 (72.3%) | 103 (83.1%) |
| Unsupported inference | 110 (22.5%) | 30 (24.2%) |
| Instruction hallucination | 58 (11.9%) | 19 (15.3%) |

**Note:** Values are presented as number of outputs with the specified failure subtype, with percentages calculated within each disease category. Individual outputs could contain more than one error or hallucination subtype; therefore, percentages do not sum to 100%.
