## Supplementary Table 6 for "Errors, Hallucinations, and Clinical Impact of General-Purpose Multimodal Large Language Models in Histopathology"

**Supplementary Table 6. Distribution of error and hallucination subtypes by disease rarity**

| **Failure subtype** | **Common**  **(n = 268 outputs)** | **Sporadic**  **(n = 252 outputs)** | **Rare**  **(n = 92 outputs)** |
| --- | --- | --- | --- |
| **Errors** |  |  |  |
| Omission | 126 (47.0%) | 148 (58.7%) | 52 (56.5%) |
| Misinterpretation | 188 (70.1%) | 180 (71.4%) | 73 (79.3%) |
| Internal inconsistency | 64 (23.9%) | 28 (11.1%) | 13 (14.1%) |
| **Hallucinations** |  |  |  |
| Fabricated feature | 184 (68.7%) | 201 (79.8%) | 71 (77.2%) |
| Unsupported inference | 61 (22.8%) | 48 (19.0%) | 31 (33.7%) |
| Instruction hallucination | 40 (14.9%) | 26 (10.3%) | 11 (12.0%) |

**Note:** Values are presented as number of outputs with the specified failure subtype, with percentages calculated within each disease-rarity category. Individual outputs could contain more than one error or hallucination subtype; therefore, percentages do not sum to 100%.
