## Supplementary Table 7 for "Errors, Hallucinations, and Clinical Impact of General-Purpose Multimodal Large Language Models in Histopathology"

**Supplementary Table 7. Distribution of error and hallucination subtypes by diagnostic efficiency**

| **Failure subtype** | **Fully sufficient**  **(n = 220outputs)** | **Additional support desirable (n = 160 outputs)** | **Additional support required (n = 232 outputs)** |
| --- | --- | --- | --- |
| **Errors** |  |  |  |
| Omission | 108 (49.1%) | 86 (53.8%) | 132 (56.9%) |
| Misinterpretation | 151 (68.6%) | 116 (72.5%) | 174 (75.0%) |
| Internal inconsistency | 26 (11.8%) | 34 (21.3%) | 45 (19.4%) |
| **Hallucinations** |  |  |  |
| Fabricated feature | 153 (69.5%) | 119 (74.4%) | 184 (79.3%) |
| Unsupported inference | 38 (17.3%) | 34 (21.3%) | 68 (29.3%) |
| Instruction hallucination | 21 (9.5%) | 21 (13.1%) | 35 (15.1%) |

**Note:** Values are presented as number of outputs with the specified failure subtype, with percentages calculated within each diagnostic-efficiency category. Individual outputs could contain more than one error or hallucination subtype; therefore, percentages do not sum to 100%.
