## Supplementary Table 8 for "Errors, Hallucinations, and Clinical Impact of General-Purpose Multimodal Large Language Models in Histopathology"

**Supplementary Table 8. Diagnostic correctness according to the presence of errors or hallucinations**

| **LLM** | **Eligible outputs, n** | **Correct without error/hallucination, n (%)** | **Correct with error/hallucination, n (%)** | **Incorrect without error/hallucination, n (%)** | **Incorrect with error/hallucination, n (%)** |
| --- | --- | --- | --- | --- | --- |
| LLM1 | 128 | 13 (10.2%) | 59 (46.1%) | 0 (0.0%) | 56 (43.8%) |
| LLM2 | 136 | 23 (16.9%) | 59 (43.4%) | 0 (0.0%) | 54 (39.7%) |
| LLM3 | 139 | 10 (7.2%) | 46 (33.1%) | 0 (0.0%) | 83 (59.7%) |
| LLM4 | 140 | 14 (10.0%) | 75 (53.6%) | 0 (0.0%) | 51 (36.4%) |
| **Overall** | **543** | **60 (11.0%)** | **239 (44.0%)** | **0 (0.0%)** | **244 (44.9%)** |

**Note:** Only outputs classified as strictly correct or incorrect were included; partially correct outputs were excluded. “With error/hallucination” indicates the presence of at least one error or at least one hallucination. “Without error/hallucination” indicates that neither an error nor a hallucination was identified. Percentages are calculated using the eligible outputs for each LLM as the denominator. LLM, large language model.
