## Supplementary Table 9 for "Errors, Hallucinations, and Clinical Impact of General-Purpose Multimodal Large Language Models in Histopathology"

**Supplementary Table 9. Prevalence of errors and hallucinations by organ**

| **Organ** | **No. of outputs** | **Any error, n (%)** | **Any hallucination, n (%)** |
| --- | --- | --- | --- |
| Adrenal | 8 | 6 (75.0%) | 5 (62.5%) |
| Breast | 36 | 31 (86.1%) | 27 (75.0%) |
| CNS | 36 | 25 (69.4%) | 18 (50.0%) |
| Colon | 44 | 33 (75.0%) | 34 (77.3%) |
| Endometrium | 36 | 32 (88.9%) | 33 (91.7%) |
| Esophagus | 40 | 37 (92.5%) | 37 (92.5%) |
| Kidney | 32 | 28 (87.5%) | 28 (87.5%) |
| Liver | 20 | 14 (70.0%) | 19 (95.0%) |
| Lung | 32 | 25 (78.1%) | 29 (90.6%) |
| Ovary | 36 | 32 (88.9%) | 23 (63.9%) |
| Pancreas | 16 | 15 (93.8%) | 16 (100.0%) |
| Prostate | 24 | 18 (75.0%) | 17 (70.8%) |
| Salivary gland | 28 | 20 (71.4%) | 21 (75.0%) |
| Skin | 48 | 41 (85.4%) | 38 (79.2%) |
| Soft tissue | 32 | 21 (65.6%) | 17 (53.1%) |
| Stomach | 44 | 35 (79.5%) | 31 (70.5%) |
| Testis | 16 | 16 (100.0%) | 16 (100.0%) |
| Thyroid | 28 | 26 (92.9%) | 17 (60.7%) |
| Urinary bladder | 24 | 19 (79.2%) | 19 (79.2%) |
| Uterine cervix | 32 | 28 (87.5%) | 24 (75.0%) |

**Note:** Values represent the number and percentage of LLM outputs containing at least one error or at least one hallucination within each organ. Because an individual output could contain both an error and a hallucination, the two categories are not mutually exclusive. Each case generated four LLM outputs. CNS, central nervous system; LLM, large language model.
