## Supplementary Table 10 for "Errors, Hallucinations, and Clinical Impact of General-Purpose Multimodal Large Language Models in Histopathology"

**Supplementary Table 10. Prevalence of errors by organ and LLM**

| **Organ** | **LLM1, n/N (%)** | **LLM2, n/N (%)** | **LLM3, n/N (%)** | **LLM4, n/N (%)** |
| --- | --- | --- | --- | --- |
| Adrenal | 1/2 (50.0%) | 2/2 (100.0%) | 1/2 (50.0%) | 2/2 (100.0%) |
| Breast | 8/9 (88.9%) | 7/9 (77.8%) | 8/9 (88.9%) | 8/9 (88.9%) |
| CNS | 7/9 (77.8%) | 5/9 (55.6%) | 6/9 (66.7%) | 7/9 (77.8%) |
| Colon | 8/11 (72.7%) | 7/11 (63.6%) | 9/11 (81.8%) | 9/11 (81.8%) |
| Endometrium | 9/9 (100.0%) | 8/9 (88.9%) | 8/9 (88.9%) | 7/9 (77.8%) |
| Esophagus | 8/10 (80.0%) | 9/10 (90.0%) | 10/10 (100.0%) | 10/10 (100.0%) |
| Kidney | 7/8 (87.5%) | 5/8 (62.5%) | 8/8 (100.0%) | 8/8 (100.0%) |
| Liver | 5/5 (100.0%) | 3/5 (60.0%) | 3/5 (60.0%) | 3/5 (60.0%) |
| Lung | 6/8 (75.0%) | 7/8 (87.5%) | 6/8 (75.0%) | 6/8 (75.0%) |
| Ovary | 9/9 (100.0%) | 6/9 (66.7%) | 9/9 (100.0%) | 8/9 (88.9%) |
| Pancreas | 4/4 (100.0%) | 4/4 (100.0%) | 4/4 (100.0%) | 3/4 (75.0%) |
| Prostate | 4/6 (66.7%) | 4/6 (66.7%) | 5/6 (83.3%) | 5/6 (83.3%) |
| Salivary gland | 4/7 (57.1%) | 4/7 (57.1%) | 7/7 (100.0%) | 5/7 (71.4%) |
| Skin | 12/12 (100.0%) | 8/12 (66.7%) | 11/12 (91.7%) | 10/12 (83.3%) |
| Soft tissue | 3/8 (37.5%) | 6/8 (75.0%) | 5/8 (62.5%) | 7/8 (87.5%) |
| Stomach | 10/11 (90.9%) | 9/11 (81.8%) | 10/11 (90.9%) | 6/11 (54.5%) |
| Testis | 4/4 (100.0%) | 4/4 (100.0%) | 4/4 (100.0%) | 4/4 (100.0%) |
| Thyroid | 7/7 (100.0%) | 6/7 (85.7%) | 7/7 (100.0%) | 6/7 (85.7%) |
| Urinary bladder | 6/6 (100.0%) | 3/6 (50.0%) | 6/6 (100.0%) | 4/6 (66.7%) |
| Uterine cervix | 7/8 (87.5%) | 7/8 (87.5%) | 8/8 (100.0%) | 6/8 (75.0%) |

**Note:** Values represent the number of outputs containing at least one error divided by the total number of cases evaluated by each LLM within that organ, with percentages in parentheses. Errors included omission, misinterpretation, and internal inconsistency. Each LLM evaluated every case. CNS, central nervous system; LLM, large language model.
