## Supplementary Table 11 for "Errors, Hallucinations, and Clinical Impact of General-Purpose Multimodal Large Language Models in Histopathology"

**Supplementary Table 11. Prevalence of hallucinations by organ and LLM**

| **Organ** | **LLM1, n/N (%)** | **LLM2, n/N (%)** | **LLM3, n/N (%)** | **LLM4, n/N (%)** |
| --- | --- | --- | --- | --- |
| Adrenal | 2/2 (100.0%) | 1/2 (50.0%) | 1/2 (50.0%) | 1/2 (50.0%) |
| Breast | 8/9 (88.9%) | 6/9 (66.7%) | 6/9 (66.7%) | 7/9 (77.8%) |
| CNS | 4/9 (44.4%) | 6/9 (66.7%) | 4/9 (44.4%) | 4/9 (44.4%) |
| Colon | 9/11 (81.8%) | 8/11 (72.7%) | 9/11 (81.8%) | 8/11 (72.7%) |
| Endometrium | 8/9 (88.9%) | 8/9 (88.9%) | 8/9 (88.9%) | 9/9 (100.0%) |
| Esophagus | 7/10 (70.0%) | 10/10 (100.0%) | 10/10 (100.0%) | 10/10 (100.0%) |
| Kidney | 7/8 (87.5%) | 6/8 (75.0%) | 7/8 (87.5%) | 8/8 (100.0%) |
| Liver | 5/5 (100.0%) | 4/5 (80.0%) | 5/5 (100.0%) | 5/5 (100.0%) |
| Lung | 7/8 (87.5%) | 7/8 (87.5%) | 7/8 (87.5%) | 8/8 (100.0%) |
| Ovary | 5/9 (55.6%) | 5/9 (55.6%) | 7/9 (77.8%) | 6/9 (66.7%) |
| Pancreas | 4/4 (100.0%) | 4/4 (100.0%) | 4/4 (100.0%) | 4/4 (100.0%) |
| Prostate | 4/6 (66.7%) | 4/6 (66.7%) | 5/6 (83.3%) | 4/6 (66.7%) |
| Salivary gland | 4/7 (57.1%) | 5/7 (71.4%) | 7/7 (100.0%) | 5/7 (71.4%) |
| Skin | 8/12 (66.7%) | 8/12 (66.7%) | 12/12 (100.0%) | 10/12 (83.3%) |
| Soft tissue | 4/8 (50.0%) | 4/8 (50.0%) | 3/8 (37.5%) | 6/8 (75.0%) |
| Stomach | 6/11 (54.5%) | 9/11 (81.8%) | 11/11 (100.0%) | 5/11 (45.5%) |
| Testis | 4/4 (100.0%) | 4/4 (100.0%) | 4/4 (100.0%) | 4/4 (100.0%) |
| Thyroid | 5/7 (71.4%) | 3/7 (42.9%) | 6/7 (85.7%) | 3/7 (42.9%) |
| Urinary bladder | 5/6 (83.3%) | 4/6 (66.7%) | 5/6 (83.3%) | 5/6 (83.3%) |
| Uterine cervix | 5/8 (62.5%) | 5/8 (62.5%) | 7/8 (87.5%) | 7/8 (87.5%) |

**Note:** Values represent the number of outputs containing at least one hallucination divided by the total number of cases evaluated by each LLM within that organ, with percentages in parentheses. Hallucinations included fabricated features, unsupported inferences, and instruction hallucinations. Each LLM evaluated every case. CNS, central nervous system; LLM, large language model.
