## Supplementary Table 12 for "Errors, Hallucinations, and Clinical Impact of General-Purpose Multimodal Large Language Models in Histopathology"

**Supplementary Table 12. Distribution of error subtypes by organ**

| **Organ** | **No. of outputs** | **Omission, n (%)** | **Misinterpretation, n (%)** | **Internal inconsistency, n (%)** |
| --- | --- | --- | --- | --- |
| Adrenal | 8 | 4 (50.0%) | 5 (62.5%) | 4 (50.0%) |
| Breast | 36 | 22 (61.1%) | 21 (58.3%) | 2 (5.6%) |
| CNS | 36 | 18 (50.0%) | 23 (63.9%) | 0 (0.0%) |
| Colon | 44 | 16 (36.4%) | 29 (65.9%) | 0 (0.0%) |
| Endometrium | 36 | 28 (77.8%) | 30 (83.3%) | 19 (52.8%) |
| Esophagus | 40 | 14 (35.0%) | 36 (90.0%) | 5 (12.5%) |
| Kidney | 32 | 23 (71.9%) | 28 (87.5%) | 3 (9.4%) |
| Liver | 20 | 6 (30.0%) | 12 (60.0%) | 0 (0.0%) |
| Lung | 32 | 9 (28.1%) | 22 (68.8%) | 1 (3.1%) |
| Ovary | 36 | 18 (50.0%) | 31 (86.1%) | 6 (16.7%) |
| Pancreas | 16 | 5 (31.2%) | 14 (87.5%) | 1 (6.2%) |
| Prostate | 24 | 16 (66.7%) | 18 (75.0%) | 0 (0.0%) |
| Salivary gland | 28 | 14 (50.0%) | 20 (71.4%) | 4 (14.3%) |
| Skin | 48 | 27 (56.2%) | 31 (64.6%) | 14 (29.2%) |
| Soft tissue | 32 | 16 (50.0%) | 21 (65.6%) | 17 (53.1%) |
| Stomach | 44 | 28 (63.6%) | 28 (63.6%) | 0 (0.0%) |
| Testis | 16 | 13 (81.2%) | 15 (93.8%) | 11 (68.8%) |
| Thyroid | 28 | 25 (89.3%) | 16 (57.1%) | 2 (7.1%) |
| Urinary bladder | 24 | 17 (70.8%) | 18 (75.0%) | 1 (4.2%) |
| Uterine cervix | 32 | 7 (21.9%) | 23 (71.9%) | 15 (46.9%) |

**Note:** Values represent the number and percentage of LLM outputs containing each error subtype within the corresponding organ. Individual outputs could contain more than one error subtype; therefore, percentages do not sum to 100%. Error subtypes were defined as omission, misinterpretation, and internal inconsistency. Each case generated four LLM outputs. CNS, central nervous system; LLM, large language model.
