## Supplementary Table 13 for "Errors, Hallucinations, and Clinical Impact of General-Purpose Multimodal Large Language Models in Histopathology"

**Supplementary Table 13. Distribution of hallucination subtypes by organ**

| **Organ** | **No. of outputs** | **Fabricated feature, n (%)** | **Unsupported inference, n (%)** | **Instruction hallucination, n (%)** |
| --- | --- | --- | --- | --- |
| Adrenal | 8 | 5 (62.5%) | 3 (37.5%) | 3 (37.5%) |
| Breast | 36 | 27 (75.0%) | 8 (22.2%) | 0 (0.0%) |
| CNS | 36 | 16 (44.4%) | 3 (8.3%) | 0 (0.0%) |
| Colon | 44 | 32 (72.7%) | 10 (22.7%) | 0 (0.0%) |
| Endometrium | 36 | 31 (86.1%) | 24 (66.7%) | 24 (66.7%) |
| Esophagus | 40 | 36 (90.0%) | 18 (45.0%) | 1 (2.5%) |
| Kidney | 32 | 27 (84.4%) | 6 (18.8%) | 17 (53.1%) |
| Liver | 20 | 19 (95.0%) | 0 (0.0%) | 0 (0.0%) |
| Lung | 32 | 29 (90.6%) | 4 (12.5%) | 0 (0.0%) |
| Ovary | 36 | 22 (61.1%) | 10 (27.8%) | 0 (0.0%) |
| Pancreas | 16 | 16 (100.0%) | 4 (25.0%) | 6 (37.5%) |
| Prostate | 24 | 17 (70.8%) | 0 (0.0%) | 3 (12.5%) |
| Salivary gland | 28 | 21 (75.0%) | 15 (53.6%) | 2 (7.1%) |
| Skin | 48 | 38 (79.2%) | 3 (6.2%) | 2 (4.2%) |
| Soft tissue | 32 | 16 (50.0%) | 0 (0.0%) | 2 (6.2%) |
| Stomach | 44 | 30 (68.2%) | 7 (15.9%) | 0 (0.0%) |
| Testis | 16 | 15 (93.8%) | 10 (62.5%) | 10 (62.5%) |
| Thyroid | 28 | 17 (60.7%) | 0 (0.0%) | 0 (0.0%) |
| Urinary bladder | 24 | 18 (75.0%) | 8 (33.3%) | 7 (29.2%) |
| Uterine cervix | 32 | 24 (75.0%) | 7 (21.9%) | 0 (0.0%) |

**Note:** Values represent the number and percentage of LLM outputs containing each hallucination subtype within the corresponding organ. Individual outputs could contain more than one hallucination subtype; therefore, percentages do not sum to 100%. Hallucination subtypes were defined as fabricated feature, unsupported inference, and instruction hallucination. Each case generated four LLM outputs. CNS, central nervous system; LLM, large language model.
