## Supplementary Table 14 for "Errors, Hallucinations, and Clinical Impact of General-Purpose Multimodal Large Language Models in Histopathology"

**Supplementary Table 14. Error and hallucination burden by organ**

| **Organ** | **No. of cases** | **Error burden, mean ± SD (95% CI)** | **Hallucination burden, mean ± SD (95% CI)** |
| --- | --- | --- | --- |
| Adrenal | 2 | 2.88 ± 2.80 (−0.42–6.17) | 2.25 ± 2.60 (−0.52–5.02) |
| Breast | 9 | 2.22 ± 1.88 (1.68–2.76) | 1.75 ± 1.57 (1.18–2.32) |
| CNS | 9 | 1.47 ± 1.36 (0.80–2.15) | 0.67 ± 0.86 (0.39–0.94) |
| Colon | 11 | 1.82 ± 1.51 (1.25–2.39) | 1.75 ± 1.48 (1.12–2.38) |
| Endometrium | 9 | 3.53 ± 2.30 (2.72–4.34) | 4.33 ± 2.80 (3.18–5.49) |
| Esophagus | 10 | 2.65 ± 1.35 (2.08–3.22) | 3.20 ± 1.67 (2.48–3.92) |
| Kidney | 8 | 3.19 ± 1.80 (2.51–3.86) | 2.94 ± 2.02 (2.15–3.73) |
| Liver | 5 | 1.45 ± 1.36 (0.69–2.21) | 2.70 ± 0.80 (2.28–3.13) |
| Lung | 8 | 1.88 ± 1.43 (1.22–2.53) | 2.25 ± 1.08 (1.74–2.76) |
| Ovary | 9 | 3.14 ± 2.34 (2.36–3.92) | 1.44 ± 1.61 (0.81–2.08) |
| Pancreas | 4 | 2.75 ± 1.81 (1.96–3.54) | 3.38 ± 1.59 (2.76–3.99) |
| Prostate | 6 | 2.67 ± 1.93 (1.64–3.70) | 1.79 ± 1.59 (0.96–2.62) |
| Salivary gland | 7 | 3.32 ± 3.02 (2.19–4.45) | 2.79 ± 2.25 (1.78–3.79) |
| Skin | 12 | 2.48 ± 1.79 (1.88–3.08) | 2.19 ± 1.36 (1.62–2.76) |
| Soft tissue | 8 | 3.88 ± 3.47 (2.88–4.87) | 0.97 ± 1.09 (0.48–1.45) |
| Stomach | 11 | 2.20 ± 1.50 (1.65–2.76) | 1.68 ± 1.36 (1.38–1.99) |
| Testis | 4 | 5.25 ± 3.17 (4.13–6.37) | 4.06 ± 2.74 (2.90–5.23) |
| Thyroid | 7 | 3.11 ± 1.79 (2.15–4.06) | 1.36 ± 1.25 (0.68–2.04) |
| Urinary bladder | 6 | 2.54 ± 1.91 (1.79–3.29) | 2.92 ± 2.34 (1.37–4.46) |
| Uterine cervix | 8 | 2.38 ± 1.79 (1.69–3.06) | 1.91 ± 1.75 (1.17–2.65) |

**Note:** Values are presented as mean ± standard deviation (SD) with 95% confidence intervals (CIs). Error burden was calculated as the summed scores for omission, misinterpretation, and internal inconsistency, and hallucination burden as the summed scores for fabricated feature, unsupported inference, and instruction hallucination, each ranging from 0 to 9 per output. Each case generated four LLM outputs. Confidence intervals were calculated from the distribution of outputs within each organ. CNS, central nervous system; CI, confidence interval; LLM, large language model; SD, standard deviation.
